# Social Learning in a Network Model of Covid-19

**DOI:** 10.1101/2020.07.30.20164855

**Authors:** Allan Davids, Gideon Du Rand, Co-Pierre Georg, Tina Koziol, Joeri Anton Schasfoort

**Affiliations:** University of Cape Town; Stellenbosch University

**Keywords:** Covid-19, Social Learning, DeGroot Model, Epidemiological Network Model, Vaccination Strategy

## Abstract

This paper studies the effects of social learning on the transmission of Covid-19 in a network model. We calibrate our model to detailed data for Cape Town, South Africa and show that the inclusion of social learning improves the prediction of excess fatalities, reducing the best-fit squared difference from 19.34 to 11.40. The inclusion of social learning both flattens and shortens the curves for infections, hospitalizations, and excess fatalities, which is qualitatively different from *flattening the curve* by reducing the contact rate or transmission probability through non-pharmaceutical interventions. While social learning reduces infections, this alone is not sufficient to curb the spread of the virus because learning is slower than the disease spreads. We use our model to study the efficacy of different vaccination strategies and find that vaccinating vulnerable groups first leads to a 72% reduction in fatalities and 5% increase in total infections compared to a random-order benchmark. By contrast, using a contact-based vaccination strategy reduces infections by only 0.9% but results in 42% more fatalities relative to the benchmark.

## 1 Introduction

While epidemiological models have been crucial in steering policy responses to the Covid-19 pandemic, their predictive performance has been poor (Ioannidis, Cripps and Tanner, 2020; Moein et al., 2021).^1^ Economists have focused their efforts to improve the predictive power of epidemiological models towards adding more realistic human behaviour.^2^ One way to do this is via social learning (Golub and Sadler, 2016). Recent empirical evidence (Bailey et al., 2021; Makridis and Wang, 2020) shows that individuals reduce their mobility more if contacts in their social networks live in areas more severely affected by Covid-19.

In Section 2, we introduce naive social learning (DeGroot, 1974) in a model of Covid-19 transmission among physically interacting agents. In our model, agents are represented as nodes in a network and their interactions as edges. Each agent has an epidemiological state corresponding to the Susceptible, Exposed, Infected, Recovered (SEIR) compartments in the tradition of Kermack and McKendrick (1927). Each day, agents change their state between these compartments. As in SEIR models, the spread of the virus—measured by the reproductive number R— is determined by two factors: (1) the contact rate, which is the frequency with which different agents interact; and (2) the transmission probability, which is the likelihood an infection takes place if two agents interact. In line with other state-of-the-art economic Covid-19 models (El-lison, 2020), our model features a heterogeneous contact rate that depends on agent age and location in the city, calibrated using demographic data on age-distributions, the composition of households, age-based contact matrices, and travel surveys. Similarly to other agent-based Covid-19 models such as Rockett et al. (2020); Almagor and Picascia (2020), and the well-known Imperial College model of Ferguson et al. (2020), the transmission probability is fixed while the contact rate can be directly influenced by the government through non-pharmaceutical interventions (NPIs), also known, and hereafter referred to as *lockdown regulations*.

However, in contrast to these models, our agents endogenously determine their contact rate based on both the government signal about the prevalence of Covid-19 as well as a social signal that is informed by the epidemiological state of connected agents. The social signal is determined by a form of naive social learning that closely follows Dasaratha, Golub and Hak (2020) and earlier work by Golub and Jackson (2010). Agents in our model not only learn from information about infections among their network contacts, but also from their neighbors’ decision to change their behavior in response to infections in their environment. Both inform an agent’s decision to physically interact with other agents, which in turn directly affects the transmission of Covid-19. The latter is a key channel of social learning in networks (Golub and Sadler, 2016), consistent with recent empirical evidence on the role of social networks during pandemics (Bailey et al., 2021).

Using this model, we study the impact of social learning on the spread of Covid-19 in Cape Town, South Africa. Cape Town is particularly interesting because all South African epidemiological models overestimated both the projected number of infections at the peak of the first wave as well as its duration—some significantly so.^3^ This was surprising since the South African government, due to economic considerations, started relaxing the stringent national lockdown policies while infections were still rising.^4^ Therefore, most models predicted an exponential increase in infections in South Africa.^5^ Despite relaxing the lockdown during this period, the peak of the excess fatality curve—generally considered the most reliable measure of Covid-19—was surprisingly low and it ended quicker than expected.^6^

To initialise our model, we use demographic data from the South African National census, contact matrices from Prem, Cook and Jit (2017), the National Household travel survey, initial detected cases from the Western Cape government, information about the government signal from the Oxford Stringency Index (Hale et al., 2020), information about observed mobility from Google, and fatality statistics recorded in South African hospitals. For epidemiological parameters we use estimates found in the extant literature. The remaining free parameters of our model, that we could not calibrate using these sources, are the transmission probability, the number of initial infections as well as two parameters related to the weight and distribution of social learning.^7^ We discuss all parameters in detail in Section 3.

Results are shown in Section 4. We, first, jointly estimate the free parameters for both a version of the model with- and one without social learning, in each version finding the parameter choices to minimize a loss function as in Nelder and Mead (1965). In the model with social learning, we obtain a best-fit squared difference between the model prediction and the empirically observed excess fatality curve of 11.40, while for the model without social learning the best-fit squared difference is 19.34. It should be noted that there are models which exhibit a better fit (Ambrosio and Aziz-Alaoui, 2020; Acuña-Zegarra, Santana-Cibrian and Velasco-Hernandez, 2020). However, in these cases a functional form is chosen for the reproductive rate as the pandemic progresses, implying that either the contact rate or transmission probability (or both) have an *exogenous* functional form. In contrast, in our model, the reproductive rate *endogenously* slows down as the virus spreads through the network, with a contact rate that is determined by the behaviour of our agents while the transmission probability is fixed.

To better understand how social learning improves the model fit over and above a calibration based on transmission probability—the standard free parameter in these types of models—we perform two sensitivity analyses, independently varying the weight of social learning and the transmission probability.

We find that a higher weight for social learning simultaneously decreases the height and duration of the curves for the infection, hospitalization, fatality, and excess fatality. For example, in our calibrated model, increasing the weight of social learning from 0.85 to 0.95 leads to 7% fewer infections and 14% fewer fatalities while keeping the peak infection day the same. In contrast, a decreased transmission probability of 2% reduces infections by 6%, fatalities by 14%. However, it simultaneously pushes the peak infection day out 3 days.

Furthermore, we find that in contrast to changing the transmission probability, the effect of changing the weight of social learning is non-linear. If we increase the strength of the social signal relative to the private signal, there is very little effect on infections for relative strengths between 0.0 and 0.8. However, between a relative strength of 0.8 and 1.0, the fatalities and infections are strongly reduced until plateauing at 76% of what infections would be without social learning. In our calibration exercise, we find a best-fit relative strength of the social signal of 0.85.

To further understand social learning, we explore how social learning affects the effectiveness of lockdown regulations by comparing the calibrated model with lockdown regulations to a hypothetical scenario in which there were no government interventions. In particular, we are interested if adding behaviour to the model would alter the conclusion that a strict lockdown was necessary, which is one of the main criticisms raised against the standard epidemiological Covid-19 models (see, for example, Shen, Taleb and Bar-Yam (2020); Squazzoni et al. (2020)). Comparing our baseline lockdown scenario to a no-intervention alternative in which agents voluntarily reduce their interactions, we find that lockdown regulations remain highly effective even when social learning is introduced.

With best-fit relative strength of social learning, lockdown regulations reduce end-of-simulation infections from 81% to 49% of the population. Furthermore, the intervention flattens the curve and pushes out the peak of infections by fives weeks. Also, under the intervention scenario, hospitals are overburdened for 12 days, rather than the 51 days as in the no-intervention scenario. As a consequence of both reduced infections and a less overburdened hospital system, total deaths are reduced by 62% in the intervention scenario, compared to the no-intervention scenario.^8^ Under the intervention scenario agents reduce their contacts on average by 50% (from 18 daily contacts to 9), while under the no-intervention scenario they reduce contacts voluntarily by 19% (from 18 daily contacts to 16).

Social learning reduces infections in the intervention scenario, and even more so in the no-intervention scenario. Introducing social learning together with a lockdown reduces total infections by 4.2%. Compared to this, adding social learning to models of Covid-19 disease transmission has a stronger effect in the no-intervention scenario, reducing infections by 6%, peak critical cases by 12.4% and deaths by 5.1%. Despite this, 80% of the population becomes infected, which can be explained by the fact that even without social learning compliance in the lockdown model is already high, averaging 82%, limiting the impact of social learning.

Finally, in Section 5 and before we conclude in Section 6, we explore how three age-based vaccination strategies affect fatalities, critical cases, and infections, had the vaccine been available at the start of the virus outbreak in Cape Town. The first strategy is the control group in which age classes are randomly given priority. The second strategy is a risk based strategy in which older agents get vaccinated first. In the third scenario, agents in age groups that have more connections are vaccinated first. Vaccinating the elderly first leads to a 72% reduction in fatalities compared to vaccinating in a random order (which is already effective, reducing fatalities relative to the no-intervention scenario by 65%). However, this comes at the cost of a 5% *increase* in total infections. On the other hand, the contact based vaccination strategy reduces infections by only 0.9%, compared to the random benchmark. However, it leads to 42% more fatalities than the benchmark random vaccination scenario.

Our paper relates to two separate strands of literature. First, we introduce social learning to the growing epidemiological literature studying the spread of Covid-19 using network models.^9^ And second, building on the literature that studies social learning in networks, we contribute to the nascent economic literature that incorporates learning into epidemiological models by studying the effect of social learning.

When it comes to epidemiological Covid-19 models, our model is most closely related to other detailed Covid-19 network models, including the Imperial College Model (Ferguson et al., 2020), which has been used to inform the Covid-19 strategy of the UK government. We also use the classic Susceptible-Infected-Recovered (SIR) Kermack and McKendrick (1927) structure that is the backbone of standard differential equation models and implement this within a Covid-19 network model.^10^ Our major contribution to the literature relative to these models is that we incorporate social learning, in which agents learn from their neighbours state rather than the global state.

Consequently, our model is also related to the large epidemiological literature which incorporates behaviour and learning into pandemic models—see Verelst, Willem and Beutels (2016) for a survey of this literature—in addition to a growing literature of economic epidemiological models (e.g. Toxvaerd (2020), Eichenbaum, Rebelo and Trabandt (2020), Krueger, Uhlig and Xie (2020), Dasaratha (2020)) which incorporates optimising behaviour. The optimizing mechanisms in these class of models typically involve agents who face a trade off between wanting to leave home to earn an income to satisfy their economic needs versus wanting to stay at home to minimize the risk and cost of infection, which is typically a function of aggregate infections. Our model is similar to these efforts in introducing behaviour to epidemiological model. However, rather than individuals optimising their behaviour using an aggregate state of infections, learning is social, and therefore local, in our model.

Finally, our paper is related to the literature on on social learning in in networks (Golub and Sadler (2016); DeGroot (1974); Golub and Jackson (2010)) with an epidemiological literature using network models to model the spread of diseases (Danon et al. (2011); Keeling and Eames (2005)). Our finding that social learning improves compliance and reduces infections is consistent with empirical evidence from recent and growing literature on the role of social networks in individual decision making and behaviour (Bailey et al., 2018), in particular, Bailey et al. (2021) and Charoenwong, Kwan and Pursiainen (2020) who find that increases in Covid-19 cases within an individuals social network leads to reduced mobility and increased compliance with stay-at-home orders. Within this literature, our paper is closely related to Makridis and Wang (2020) who study the impact of social learning on consumption during a pandemic, within a consumption and savings network-based model. The authors show that when compared to a model without social learning, including social learning leads to a greater reduction in consumption, as learning leads to an increased in perceived infections by agents as they internalize the infectious state of the nodes in their network. The authors focus explicitly on the role of social learning on consumption during a pandemic and do not allow for social learning to affect the progression of the disease. In that regard, our model contributes to this literature by studying how social learning affects disease progression. We also distinguish our work through how we allow social learning to influence behaviour. In Makridis and Wang (2020), agents learn about the true state of infections solely through the infection state of the nodes in their network, in the vein of Golub and Jackson (2010). On the other hand, in our model, we closely follow the implementation of social learning in Dasaratha, Golub and Hak (2020), whereby agents receive a noisy private signal of the state of the world and use a weighted combination of this noisy private signal and a social signal, derived from social learning, to form a compliance decision.

## 2 The Model

We simulate the spread of Covid-19 in a network of physical interactions modelled as an undirected graph *𝒢* = (*𝒩, ℰ*) where the set of nodes *𝒩* represent agents and the set of edges *ℰ* ⊂ (*𝒩* × *𝒩*) represent physical interactions between them.^11^

In this section, we discuss: (i) the agents and their characteristics, (ii) the network that governs interactions among agents, (iii) the epidemiological status updates and disease transmission, and (iv) the technical details of how we implement the model numerically.^12^

### 2.1 Agents

There is a set of agents *𝒩*, living in households *ℋ* in a city with districts *𝒲*.^13^ Agents *j* ∈ *𝒩* are characterized by their epidemiological state *P*_*j*_ and four agent-specific parameters, age *a*_*j*_ ∈ *𝒜*, home district *w*_*j*_ ∈ *𝒲*, district to travel to 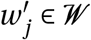 and household *h*_*j*_ ∈ *ℋ* that remain constant throughout the simulation. We model nine age groups, *𝒜* = {0−10, 10−20,…, 70−80, 80+}, and, in our specific application to the cities of Cape Town, 116 districts *𝒲*.

An agent’s epidemiological status *P*_*j*_ can take seven values: Susceptible to infection (*S*); Exposed but not infectious (*E*); Infectious but asymptomatic (*I* ^*as*^); Infectious and symptomatic (*I* ^*s*^); Critically ill (*C*); Recovered (*R*); or Deceased (*D*).^14^ The two sub-categories of the infectious status are important as there are clear indications that some individuals with Covid-19 never show symptoms but still infect others. The critically ill category is used to compare the number who require hospitalization to the critical care capacity of the health system–in our model, the probability of death increases if there are more critically ill patients than hospitals have capacity. While such agents are medically still infectious, we assume they are isolated in hospital where they are unable to infect other agents.^15^

When initialising the agents *j* ∈ *𝒩*, we assign them a district to live in *w*_*j*_ proportional to the relative share of people living in this district relative to the full city population. For each agent *j* ∈ *𝒩*_*w*_ ⊂ *𝒩*, we randomly assign an age group *a*_*j*_ using the observed district level age distribution data *F* ^age^.

### 2.2 Agent Interactions

Agents interact in the network *𝒢* that represents the possible interactions of individuals within a city, calibrated to observed contact matrices from Prem, Cook and Jit (2017), which we denote *F* ^hc^.^16^ We model agent interaction with other agents separately for household and non-household interactions.

#### 2.2.1 Household Interactions

The set of agents in district *w*, denoted *𝒩*_*w*_, is partitioned into a disjoint set of households *ℋ*_*w*_ of different sizes. Let *𝒩*_*h,w*_ be the set of agents in household *h* in district *w* (i.e.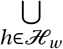 *𝒩*_*h,w*_ =*𝒩*_*w*_) with household size *N*_*h,w*_ = |*𝒩*_*h,w*_ |. Agents in each household are all connected with one another. The set of households in each district, *ℋ*_*w*_, is constructed iteratively, by randomly drawing households sizes *N*_*h,w*_ from the empirical observed distribution of households sizes specific to each district until 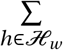 *N*_*h,w*_ = *N*_*w*_.^17^ This yields, for each district *w*, a set of house-holds *ℋ*_*w*_ of different sizes so that there are a total of *H*_*w*_ = |*ℋ*_*w*_ | households in district *w*, representative of the observed household size distribution for each district.

After initializing a realistic district level distribution of households of varying sizes, we then proceed to assign each agent *j* ∈ *𝒩* to a household *ℋ*_*w*_ in order to match the observed within-household age probability distribution *F* ^hc^. We proceed in two steps.

First, for each household *h* in district *w*, we select a household head as follows: From the set of agents in the district, we randomly select an agent 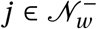 (sequentially over households, without replacement) to be head of household *h*, where 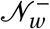 denotes the subset of all agents in *𝒩*_*w*_ unassigned to a household. The selected household head has age-category *a*_*j*_, which was assigned across agents, by district to match available data on age-distributions per district (as described in Section 2.1). This process continues until each of the *H*_*w*_ households in region *w* has a household head.

Second, for household *h* with household head of age *a*_*h*_ and household size *N*_*h,w*_, we select additional household members sequentially, randomly, and without replacement from 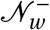 according to the probability distribution constructed from the empirical household contact matrix *F* ^hc^until there are *N*_*h,w*_ members, then move on to the next household.

#### 2.2.2 Non-Household Interactions

Non-household interactions take place in the model each day of the simulation *t* ∈ [0, *T*] when agents travel across the city. When this happens, agent *j* in district *w* belonging to household *h* forms non-household edges with agents in *𝒩* \ *𝒩*_*h,w*_. The formation of edges is constructed to be consistent with two sources of data: a non-household contact matrix from Prem, Cook and Jit (2017) which we denote *F* ^oc^and an observed travel matrix constructed using data from a nationally representative travel survey, which we denote *F* ^tv^. *F* ^oc^records the best estimates of data on daily number of contacts between individuals in different age categories *outside* of the household.^18^ *F* ^tv^records the probability that an agent in district *w* travels to district *w′* on any given day.^19^ The number of non-household connections that agent *j* makes, 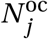 is the total number of average non-household contacts for an agent in age category *a*_*j*_ as recorded in *F* ^oc^.

The set of agents with whom agent *j* forms non-household edges is constructed in two steps.

First, agent *j* selects a *destination* district *w′* according to the empirical probabilities in *F* ^tv^. Iterating over all agents, this yields a *population of potential contacts* in each district that consists of all agents from other districts who selected it as a destination. Denote the population of potential contacts in district *w′* as 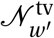. Second, for each agent *j* with destination district *w′*, we create edges to agents that are randomly selected from 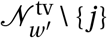, without replacement, according to the probability distribution implied by the non-household contact matrix *F* ^oc^, until 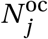 new edges have been created.^20^

To summarize, the algorithm generates the network *𝒢* which contains, for each agent *j* a neighbourhood of all contacts *𝒩*_*j*_ ⊂ *𝒩* to or from whom the virus can be transmitted. Let agent *j* be in household *h*, then *𝒩*_*j*_ is composed of two components that can be differently impacted by lockdown regulations: a neighbourhood of household contacts 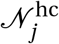 and a neighbourhood of non-household contacts 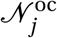, i.e. 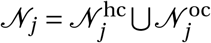.

We present a stylized network for a model with 40 agents in two districts in Figure 1 where household edges are dotted and other edges are continuous. The nodes are agents and the colours mark differences in their epidemiological status *p*_*j*_. The main clusters that can be observed represent agents that travel to the same location every day. The virus can then spread to other districts via the household links when agents travel back to their home district.

**Figure 1:**
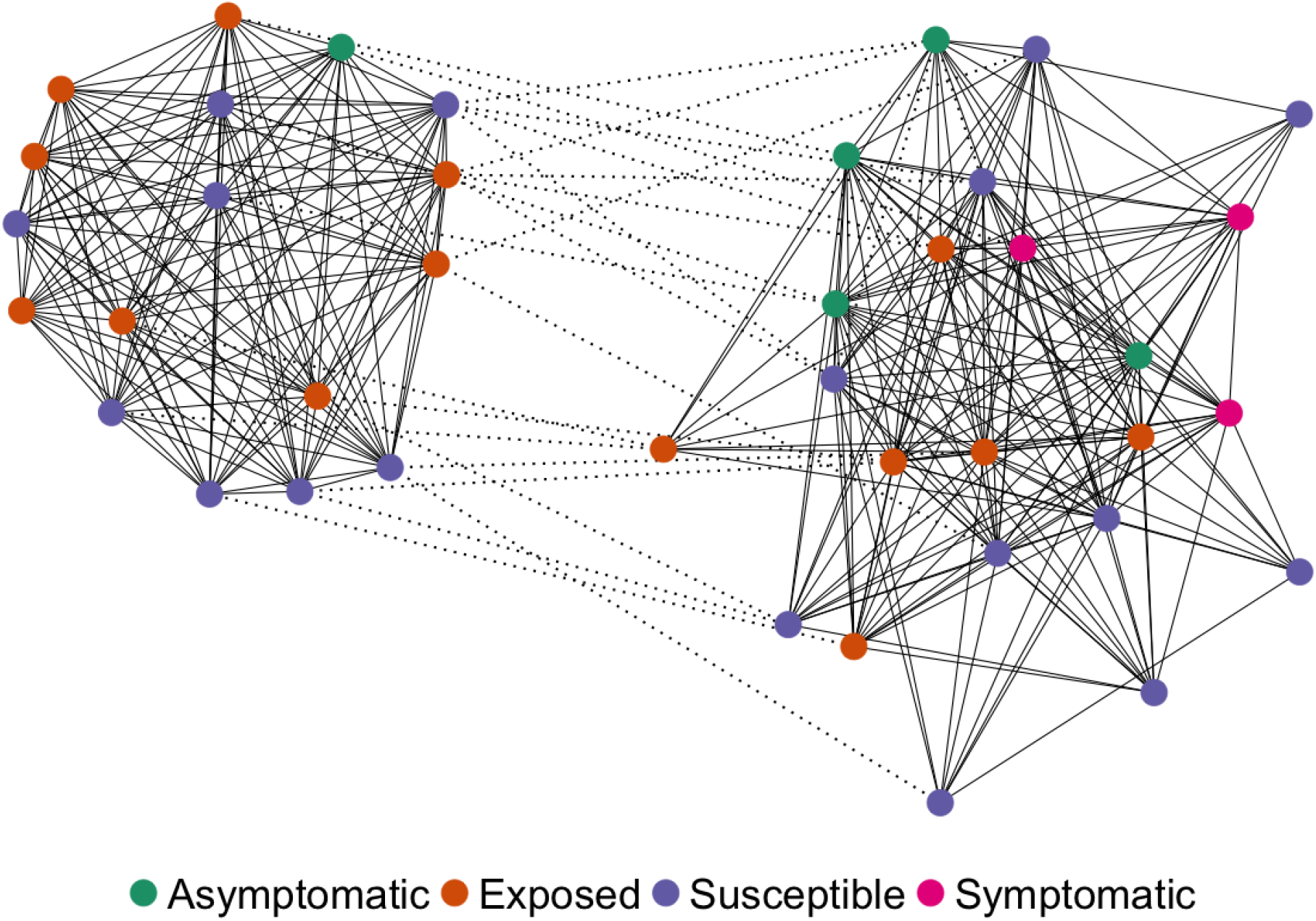
City network example. This schematic shows a 40-agent, 2-district city network at *t* = 14 generated by our algorithm. Household edges are dotted and other edges are continuous. The nodes are agents and the colours mark differences in their epidemiological status *p*_*j*_.

### 2.3 Epidemiological Status Updates and Disease Transmission

In each period of the simulation *t* ∈ [0, *T*], the epidemiological status of each agent (*P* _*j*_ (*t*)) is updated. Disease transmission takes place as agents interact. Interactions are contacts between two agents who share an edge in *E*. When a susceptible agent *i* (*P*_*i*_ (*t*) = *S*) interacts with an infectious agent *j* (*P* _*j*_ (*t*) ∈ {*I* ^as^, *I* ^s^}), transmission of the virus occurs with probability *π*^*E*^.^21^ Importantly, *π*^*E*^ is only positive for infectious agents (*P* _*j*_ (*t*) ∈ {*I* ^as^, *I* ^s^}) and is 0 otherwise. Put differently susceptible, exposed, recovered, critical and dead agents cannot infect susceptible agents. While critical agents are still infectious, we assume they are isolated in hospital where they are unable to infect other agents.

#### 2.3.1 Initial Infections

The first agents to update their epidemiological status from susceptible *S* to one of the infected compartments (*E, I* ^as^ or *I* ^s^) are agents that are created at the start of the simulation (*t* = 0). We assign initial infections based of the observed number of cases per district at the beginning of the Covid-19 outbreak, *F* ^ca^. Within each district, we pick a random agent and update their initial status to *P*_*j*_ (0) ∈ {*E, I* ^as^, *I* ^s^} (with equal probability), and assume that the agent has been in this for a random number of days 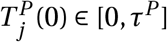, where *τ*^*P*^ is the tenure in compartment *P*.

#### 2.3.2 Evolution of Previously Infected Agents

Agents transition through the various phases or compartments of the disease via a system of between-state probabilities, which denote the probability that an agent moves from any given state to the following state. We present the disease progression schematically in Figure 2. We calibrate these probabilities using estimates from the literature which we discuss in a later section.^22^

**Figure 2:**
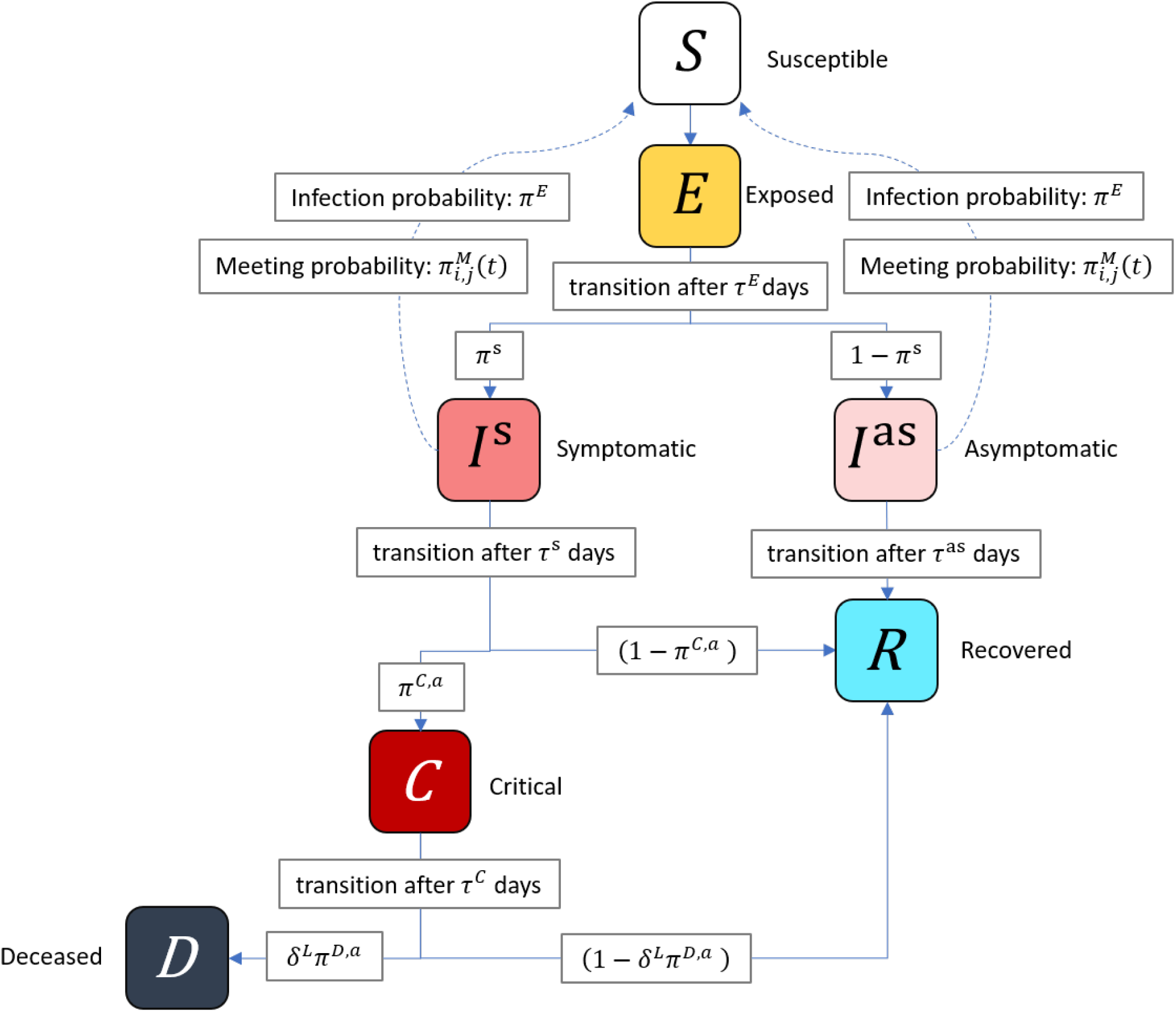
Schematic of the disease progression. This schematic shows the disease progression modelled in SABCoM. Solid arrow between compartments represent transitions of an agent to different disease statuses. Dashed arrows represent social connections along which the virus may be transmitted from infectious to susceptible individuals.

While the probability of infection *π*^*E*^, the probability of developing symptomatic version of the disease *π*^s^ are age-invariant probabilities, we model the probability of becoming critical 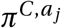 and the probability of death 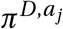 as age-varying probabilities consistent with existing evidence of varying mortality rates by age-group (Verity et al., 2020). Lastly, we augment to the probability of death 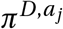 to include an important feature of the Covid-19 pandemic, increased mortality rates when the health system is overburdened. We model this via a multiplier *δ*^*L*^. We model *δ*^*L*^ as:

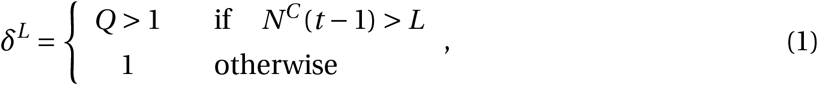

where *L* is the capacity of the health system, *N*^*C*^ (*t* − 1) is the total number of agents in the critical state in period *t* − 1, and *Q* is the empirical multiplier that increases the probability of dying when hospital capacity is overwhelmed. This generates a probability of death of 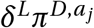.

#### 2.3.3 Agent Behaviour, Lockdown Regulations and New Infections

We study the impact that behavioural responses to lockdown regulations have on the transmission of the disease. While these interventions take many forms, such as stay-at-home orders, limits on indoor activities, and policies that encourage proper hygiene, mask wearing and social distancing. For simplicity, we consider only one aspect of lockdown policies in our model: those that reduce physical interaction among agents.

The major methodological contribution of this paper is to consider how social learning affects the compliance of individuals with lockdown regulations and in turn study how this affects disease transmission. Our implementation of social learning closely follows that of Dasaratha, Golub and Hak (2020).

In each period *t*, every agent *j* observes a public signal *ξ*(*t*) and a private signal *ζ*_*j*_ (*t*). The public signal represents the stringency of lockdown measures put in place. We model the public signal using a time varying lockdown stringency index 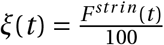 from Hale et al. (2020), measuring the stringency of the lockdown in South Africa relative to all other countries, where 1 represents the most stringent lockdown and 0 represents the least stringent. Each agent then observes a unique private signal *ζ*_*j*_ (*t*) which consists of the public signal *ξ*(*t*) and an individual noise term *ε*_*j*_ (*t*), where *ε*_*j*_ (*t*) is drawn independently for each *j* from a truncated normal distribution with support [−*ξ*(*t*), 1 − *ξ*(*t*)] and mean 0 and standard deviation *σ*, such that *ζ*_*j*_ (*t*) ∈ [0, 1]. Formally,

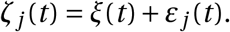

After observing their private signal, agents proceed to set their level of compliance *ϕ*_*j*_ (*t*) with policies that aim to reduce physical interaction. We model the level of compliance *ϕ*_*j*_ (*t*) of each agent as a weighted average of the private signal *ζ*_*j*_ (*t*) (with weight of *ρ*) and a social signal (with weight of (1 − *ρ*)). The social signal is the simple average of observed *t* − 1 stringency of compliance of all neighbours. Formally 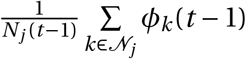:

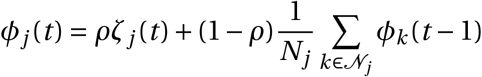

In this way, compliance depends both on an agents observation of the aggregate stringency of lockdown measures as well as the compliance levels of their neighbours. After choosing a level of compliance, a physical meeting between two agents *i* and *j* occurs with a time-varying, pair-specific probability 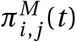 modelled as:

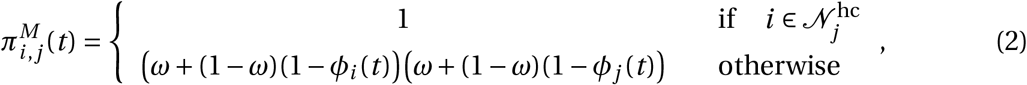

The probability 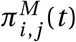 firstly depends on nature of contacts - whether these contacts occur within the household or outside the household. Agent *j* has daily physical meetings with all members of their household, so that if 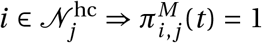. Second, for members in the set of non-household contacts of agent *j*, the probability of a meeting between *j* and 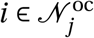 depends on three parameters, *ω* ∈ [0, 1] and *ϕ*_*i*_ (*t*) and *ϕ*_*j*_ (*t*).

We model *ω* ∈ [0, 1] to represent the minimum share of contacts that each agent will still visit under the most stringent lockdown regulations, where *ω* = 0 means that under the strictest lockdown an agent will have no contacts whereas *ω* = 1 means that even if agents fully comply with the strictest possible lockdown agents will have full contacts. Therefore, in between these two extremes, this parameter reflects the fact that agents will still have some contacts even under a very strict lockdown. After all, while lockdowns restrict movement, many agents will still need to leave home, most notably essential workers.

The probability that agent *i* and agent *j* meet is therefore a function of their own adherence to lockdown regulations and the aggregate reduction in mobility. Put differently, when governments enact a lockdown, the restrictions in place limit full mobility, however, some mobility always occurs (*ω*). Over and above this level of minimal mobility, agents can make additional contacts, and the extent to which they make additional contacts, depends on their non-compliance with regulation (1 − *ϕ*). To build intuition consider the extreme case where *ϕ*_*i*_ (*t*) = 1 and *ϕ*_*j*_ (*t*) = 1 and agents fully comply. The probability of meeting reduces to *ω*^2^, the minimum amount of mobility.^23^ This functional form therefore encodes two features: firstly even with full compliance to policy, some physical meetings may happen during the course of everyday life, and (ii) individuals can always coordinate to ensure a meeting should they wish to do so strongly enough. An alternative way of thinking about this, within our model framework, is that without any intervention the base assumption is that individuals meet with their non-household contacts with certainty every day. Without the inclusion of *ω*, the probability of two agents meeting would reduce to a function of their non-compliance, and two fully compliant agents would never meet. Given what we know about essential workers, contacts do still occur, even in a full lockdown. The inclusion of *ω* therefore prevents agents from being able to fully cut themselves off from making connections in their network of non-household contacts.^24^

In summary, any given period agent *i* and *j* physically meets with probability 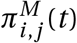. Conditional on a meeting, if one agent is susceptible and the other is infectious, the virus is transmitted with probability *π*^*E*^.

### 2.4 Implementation

We implement the model using Python using publicly available data, ensuring full reproducibility.^25^ In addition to the variables defined above, we make use of auxiliary (internal) variables during the simulation to keep track of how long each agent has been in each epidemiological state. Furthermore, we keep track of the number of other agents have been infected by each agent in the current period as well as in total. Finally, we store each agent’s neighbours, their number of contacts, as well as the household number neighbors belong to.

For our first simulation day, we record the sets of nodes *𝒩* and edges *ℰ*. On every simulated day *t*, we keep track of the epidemiological status *P*, district number *w*_*j*_, and age group *a*_*j*_ of every agent along with the number of other agents it has infected during that day. This data is recorded and stored.

We use Monte Carlo methods, simulating the model *V* times. For each simulation, we seed the pseudo random number generator with a different number *v* ∈ *𝒱*, ensuring both that its results can be replicated and that changes in output for the same seed can be attributed to changes in policy rather than changes in stochastic factors.

## 3 Calibration

Our model calibration can be divided into four parts. First, we set a baseline for the simulation-specific parameters. Second, we assign values to parameters that are associated with the clinical features of Covid-19 progression. Third, we calibrate the geo-spatial and demographic features to data for Cape Town. Finally, we estimate the remaining parameters for which we could not find a value in the data or literature.

Overall, there are 16 parameters and 9 input files used in the numerical simulation. Of the parameters, 3 are simulation parameters, 9 are related to the Covid-19 pathogen (Table 1), 2 are related to social learning, 1 considers the capacity of the health system, and 1 is related to the effectiveness of lockdown policies. In this section, we discuss how we calibrate these parameters. The values have been set using data and literature wherever possible. However, there are four parameters for which we were not able to find values in the data or literature. Therefore, we estimate them based on excess fatality data.

**Table 1:** Covid-19 pathogen related parameters This table reports parameter choices for the Covid-19 pathogen used in the model. For each parameter, we report the chosen value and the applicable medical study.

| Name | Description | Value | Source(s) |
| --- | --- | --- | --- |
| Latency period ($\tau^E$) | Length of incubation phase | 4 | Silal et al. (2020) |
| Recovery period asymptomatic infection ($\tau^{as}$) | Average number of days a person remains infectious if asymptomatic | 7 | Silal et al. (2020) |
| Transition period symptomatic infection ($\tau^s$) | Transition period symptomatic infection | 7 | Huang et al. (2020); Silal et al. (2020) |
| Days in Critical compartment ($\tau^C$) | Average number of days admitted to the ICU unit | 11 | Chen et al. (2020); Silal et al. (2020) |
| Probability to become Asymptomatic ($\pi^{as}$) | Probability to become an asymptomatic carrier | 0.6165 | Ing, Cocks and Green (2020)<br>Yang, Gui and Xiong (2020) |
| Probability to become critically ill ($\pi^{C,a}$)<br>(age dependent) | Probability to become critically ill and require hospitalization | PANEL B | Verity et al. (2020) |
| Probability to die ($\pi^{D,a}$)<br>(age dependent) | Probability to enter deceased state when critically ill | PANEL B | NICD |
| Health system overburdened multiplier ($\delta_L$) | Factor by which the probability to die increases if health system is over capacity | 1.79 | Chen et al. (2020) |

(a) Covid-19 pathogen related parameters
|  | 0-10 | 10-20 | 20-30 | 30-40 | 40-50 | 50-60 | 60-70 | 70-80 | 80+ |
| --- | --- | --- | --- | --- | --- | --- | --- | --- | --- |
| $\pi^{C,a}$ | 0.1% | 0.3% | 1.2% | 3.2% | 4.9% | 10.2% | 16.6% | 24.4% | 27.3% |
| $\pi^{D,a}$ | 2.1% | 3.3% | 3.4% | 5.3% | 9.7% | 15.5% | 24.9% | 30.1% | 37.1% |

(b) Age specific probabilities to enter critical or deceased states

### 3.1 Simulation Parameters

We run our simulations with *N* = 100, 000 agents. Every simulation runs for a minimum of *T* = 1, 171 days and is typically repeated *V* = 50 times using Monte Carlo simulation methods to average out stochastic effects.^26^

### 3.2 Covid-19 Parameters

With respect to parameters which pertain to the Covid-19 pathogen, we choose best available estimates from peer-reviewed medical journals. Despite there still being considerable uncertainty around the clinical course and transmission of the disease, we were able to find literature estimates for most parameters.

The Covid-19 parameters are chosen from recent studies (see Chen et al., 2020; Verity et al., 2020; Huang et al., 2020, among others). Table 1 provides an overview of all parameters and their sources.

Because we could not find a literature estimate for *π*^*E*^, the probability of transmission when an infectious agents comes into contact with a susceptible agent, we will estimate its value along with our policy parameters.^27^ This procedure will be described in section 3.4.

### 3.3 Applying the Model to Cape Town

We calibrate our model to Cape Town, which covers over 2,400 square kilometers and has a population of 3, 740, 026.^28^ The city are sub-divided in *W* = 116 administrative districts, known as wards.^29^ Calibrating our model to Cape Town means that we populate our input files with data that is representative of the city and set our health system parameter based on actual hospital capacities. In this way, our model can easily be applied to any other city, where similar data is available.

Our first input data file *F* ^pop^contains the population and age distribution per ward, The second file *F* ^hs^is the empirically observed household size distribution for all wards in the city. For each of these files, we use ward-level data from the South African National Census for 2011.

Then, we make use of social contact matrices for South Africa as a proxy for social contact matrices in Cape Town, obtained from Prem, Cook and Jit (2017). The contact matrices are the *A* × *A* age-group household contact matrix *F* ^hc^, and non-household contact matrix *F* ^oc^, respectively. These matrices specify how many average daily contacts people in a particular age group have with others, by age group. The matrices include total contacts, household contacts, as well as work and school contacts. Since we only consider household and non-household contacts, their reported household contacts form *F* ^hc^. To obtain a measure for non-household contacts, *F* ^oc^, we simply subtract household contacts, *F* ^hc^ from total contacts. We illustrate the total household and non-household social contact matrix in Table 2. Between the age groups 0-10 and 40-50 we see a significant share of interactions occur on the diagonal, indicating predominantly within age-group contacts. However, for age groups 50-60 and above, the diagonals become less significant, indicating substantial cross-age group interaction. This is a particularly important feature given mortality rates are highest among the elderly, who also have the highest cross-age group interactions, especially with the young.

**Table 2:** Age-based social contact matrix This table shows the age-based social contact matrix of total contacts for South Africa obtained from Prem, Cook and Jit (2017). In this representation, we normalize contacts row-wise within age group so each value can be interpreted as a percentage of all contacts for that specific age-group. Values should be read across columns with each column representing the person from which the contact originates and each row representing the person receiving the contact. We add one additional column and row to this table corresponding to individuals older than 80 years old. While our model contains individuals in this age group, the data we use does not include this age group. We set the contact matrix values of individuals above the age of 80 equal to the contact matrix values of individuals between 70 and 80 and therefore explicitly assume an identical contact structure.

|  | 0 - 10 | 10 - 20 | 20 - 30 | 30 - 40 | 40 - 50 | 50 - 60 | 60 - 70 | 70 - 80 | 80 + |
| --- | --- | --- | --- | --- | --- | --- | --- | --- | --- |
| 0 - 10 | 0.51 | 0.11 | 0.08 | 0.17 | 0.13 | 0.16 | 0.19 | 0.17 | 0.17 |
| 10 - 20 | 0.12 | 0.61 | 0.16 | 0.12 | 0.20 | 0.19 | 0.14 | 0.25 | 0.25 |
| 20 - 30 | 0.11 | 0.10 | 0.43 | 0.19 | 0.15 | 0.18 | 0.14 | 0.08 | 0.08 |
| 30 - 40 | 0.14 | 0.07 | 0.16 | 0.27 | 0.20 | 0.16 | 0.19 | 0.11 | 0.11 |
| 40 - 50 | 0.06 | 0.07 | 0.10 | 0.15 | 0.21 | 0.16 | 0.14 | 0.13 | 0.13 |
| 50 - 60 | 0.03 | 0.02 | 0.05 | 0.06 | 0.07 | 0.11 | 0.10 | 0.09 | 0.09 |
| 60 - 70 | 0.02 | 0.01 | 0.01 | 0.02 | 0.02 | 0.03 | 0.07 | 0.07 | 0.07 |
| 70 - 80 | 0.01 | 0.00 | 0.00 | 0.00 | 0.01 | 0.01 | 0.02 | 0.05 | 0.05 |
| 80 + | 0.01 | 0.00 | 0.00 | 0.00 | 0.01 | 0.01 | 0.02 | 0.05 | 0.05 |

Next, we construct the travel matrix *F* ^tv^using travel patterns from the 2013 National Household Travel Survey, a nationally representative travel survey undertaken by Statistics South Africa. We use these data to calculate a travel matrix which contains the probability of travel across all pairs of wards in our model, as well as the probability that an agent does not travel across wards (an example being an agent who lives and works in the same ward).^30^ We describe the process by which we map the travel survey data to our model in more detail in Appendix E.

For the distribution of initial infections, we make use of *F* ^ca^, a data set that contains the total number of detected cases per ward. For each Ward, we divide the number observed infections over the total number of infections in the metropolitan area and use this to distribute initial infections over the city. However, because detection of cases in South Africa was initially severely limited by testing capacity, we do not use detected cases to initialise infections in the simulation. Instead, we treat this parameter as uncertain and estimate its value.

We calibrate the health system capacity as follows. According to official sources, there were 2162 acute beds in the Western Cape province on the 22nd of May 2020.^31^ Additional 1428 care beds were scheduled to be provided by temporary hospitals. Of these, 89% will be in Cape Town. Assuming that this ratio holds for all beds, we set our health systems capacity to be *L*_*ct*_ = 0.000917, the fraction of acute beds available in Cape Town divided by the total population.

Finally, we calibrate *ω* ∈ [0, 1], the minimum share of contacts that agents need to have even under a full lockdown, using Google observed mobility data.^32^ We take the value of the mobility index on 26 March 2020, the day upon which South African entered into a very strict lockdown in which the streets were almost empty.^33^ We therefore assume that any travel recorded on this day, must have represented travel by essential workers. This results in *ω* = 0.46.

For the government signal, we use the Stringency Index *F* ^*str in*^ (Hale et al., 2020), published by the Oxford Blavatnik School of Government. The index scores lockdowns worldwide on a 0-100 scale where 0 is the least and 100 is the most stringent lockdown. In response to the Covid-19 outbreak, South Africa went into a 87.96 stringency index lockdown that at midnight on 26 March 2020. The stringency of this lockdown was eased to 84.26 on the first of May, to 80.56 on the first of June, and then to 76.85 on June 8^th^.

### 3.4 Estimating Uncertain Parameters

There are four parameters for which we could not find reliable values in the literature or data: the standard deviation of the shock to the private signal *σ* ∈ (0.0001, 0.1), the weight of the private signal *ρ* ∈ (0.05, 0.7), the probability of transmission *π*^*E*^ ∈ (0.025, 0.35), and initial infections *E* + *I* ^as^ + *I* ^s^ =∈ (1000, 3500). We proceed to estimate these parameters.

Since the possible number of combination of four uncertain parameters is large, we first use Latin Hypercube Sampling (Stein, 1987) to efficiently select 5 initial four-parameter combinations. These values will serve as the starting point for our estimation procedure.

Next, we take a two-step approach in which we estimate the values of all parameters. For each initial set of our parameters, we use the Simulated Method of Moments (SMM) methodology (Franke and Westerhoff, 2012).^34^ The method requires that one chooses which moments of the empirical data one wants to replicate. In our case, we decided to focus on the observed excess fatality data for the Cape Town metropolitan area over a period of 117 days, the length of *F* ^*str in*^. Following this approach, we minimise a quadratic loss function *B* using a constrained Nelder-Mead simplex algorithm (Nelder and Mead, 1965) with 10 iterations, where *B* is defined as:^35^

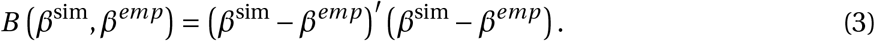

Here, *β*^*emp*^ and *β*^*sim*^ are sets containing the empirical and model-generated moments respectively. For each iteration in the optimiser, we simulate *V* = 15 Monte Carlo Simulations and score each simulation independently. We then average these scores for a final score.

Using this procedure, we jointly estimate all four uncertain parameters for all 5 initial parameters sets, pick the best fit, and then use some manual calibration for the last mile. This yields the following estimates: *σ* = 0.05, *ρ* = 0.15, *π*^*E*^ = 0.02989 and *E* + *I* ^as^ + *I* ^s^ = 928 initially infected agents, where initial infections are randomly distributed across these three compartments. This last estimate translates to 34,707 initial infections, given that our 100.000 agents represent 3740000 people.

### 3.5 Model Validation

To validate our model against data, we choose to compare simulated fatalities with realized excess fatalities in Cape Town. Excess deaths represent the number of weekly deaths recorded during 2020 relative to the average number of deaths recorded during the same weeks in 2018 and 2019.^36^ Excess deaths are regarded as the most accurate measure of COVID deaths, given concerns regarding an under-reporting of true COVID deaths in national statistics. We report the number of estimated deaths produced by our model compared to the number of excess deaths reported for Cape Town in Panel (a) of Figure 3. The root mean squared error (RMSE) between simulated and excess deaths is 11.4. Our model is able to replicate the pattern of excess deaths for Cape Town reasonably well, which provides reassurance that our model is able to capture the basic dynamics of COVID-19.

**Figure 3:**
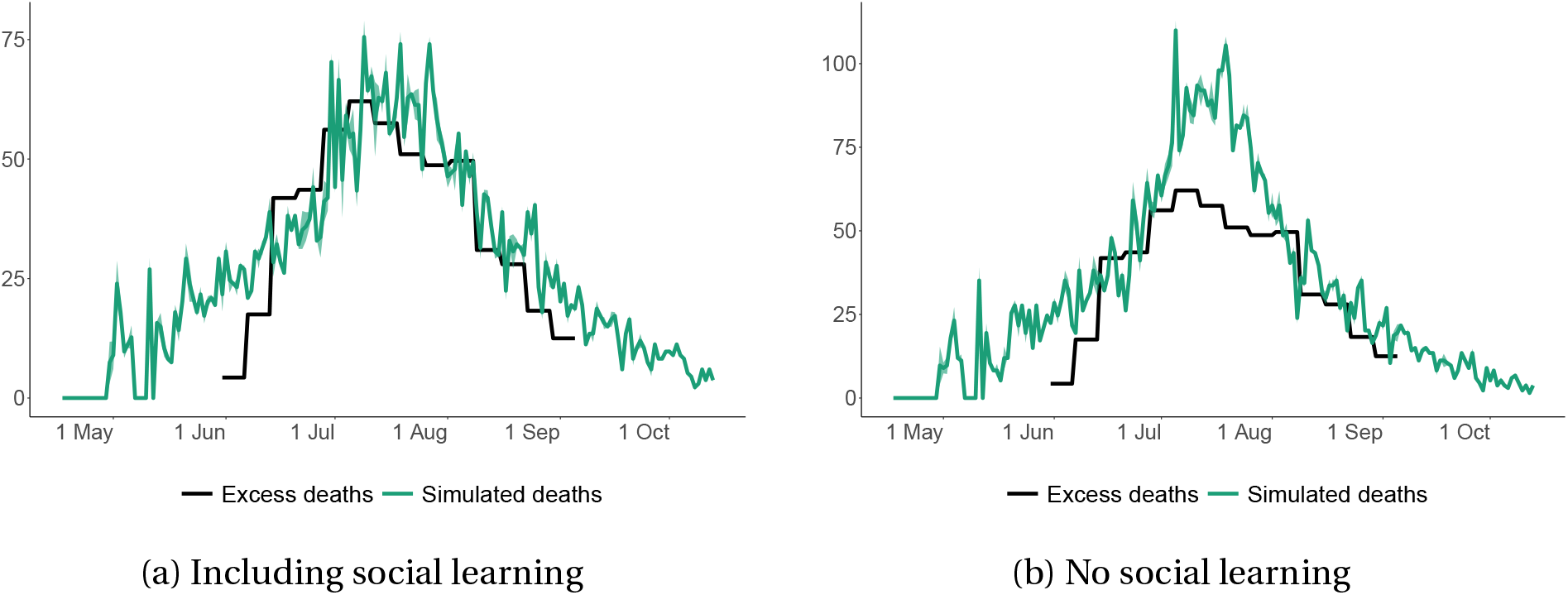
Simulated deaths and observed excess deaths with and without social learning. This figure shows how simulated deaths compares with excess deaths recorded in Cape Town in a model with social learning and a model without social learning. Both scenarios represent the best fit obtained using the calibration procedure outlined in Section 3.4. In the model including social learning, the root mean squared error (RMSE) between simulated and excess deaths is 11.40. The RMSE in the model without social learning is 19.34.

## 4 Results

In this section we present our simulation results with the goal of illustrating the role of social learning in a epidemiological Covid-19 network model. Throughout this section, we present results for simulations with our baseline model over M = 50 random seeds, reporting the average effects along with 95% confidence intervals.^37^

This section is structured as follows. First, we show the best fit we could find for the model without learning and compare it to the fit with learning. Then, we show how differences in the weight of social learning affect the spread of the virus and contrast this to the impact of differences in transmission probability. Finally, we show how lockdown policies interact with social learning.

### 4.1 Fitting Excess Death Curves With- and Without Social Learning

First, we fit the empirically observed excess fatality curve in Cape Town without social learning. Figure 3 compares the fit of a model with social learning to one without social learning. We show results for *best* fit obtained using the calibration procedure outlined in Section 3.4.

### 4.2 Social Learning, Transmission Probability, and Contact Rates

Our fitting exercise in Section 4.1 shows that social learning can improve the model fit significantly. In this section, we vary the strength of social learning and compare the effect of social learning on the curves for our main observables to the effects of varying the transmission probability and the contact rate, the other free model parameters.

Specifically, we first check how sensitive our model is to a variation of the weight of the private signal *ρ* (conversely, changing the weight of social learning, (1 − *ρ*)) around its calibrated value of *ρ* = 0.15 from *ρ* = 0.0 (only social learning) to *ρ* = 0.3 in steps of 0.05. Second, we vary the transmission probability *π*^*E*^ around its calibrated value *π*^*E*^ = 0.02989 from *π*^*E*^ = 0.02689 to *π*^*E*^ = 0.03289 in steps of 0.01. Finally, we vary the minimum share of contacts that each agent will still visit under the most stringent lockdown regulations *ω* around its calibrated value *ω* = 0.407 from *ω* = 0.257 to *ω* = 0.557 in steps of 0.05. Here, we note that, while the contact rate is endogenous in our model, *ω* affects the contact rate at every point in the simulation independently from lockdown policies and learning. Therefore, it allows us to test the broader sensitivity of the contact rate.

Figure 4 shows how these three variables impact the three most important epidemiological curves: the number of expected infections, hospitalized agents, and fatalities. It shows that while both a reduction in the weight of the private signal and a reduction in the transmission probability reduce the total number of infections, fatalities, and critical cases, social learning brings the peak of the curve forward while not making it longer whereas both a reduction in transmission probability and general contact rate flattens the curve. On the other hand, decreasing the weight of the private signal (and increasing the weight of the social signal) reduces the number of infections, but does not change the duration of the pandemic.

**Figure 4:**
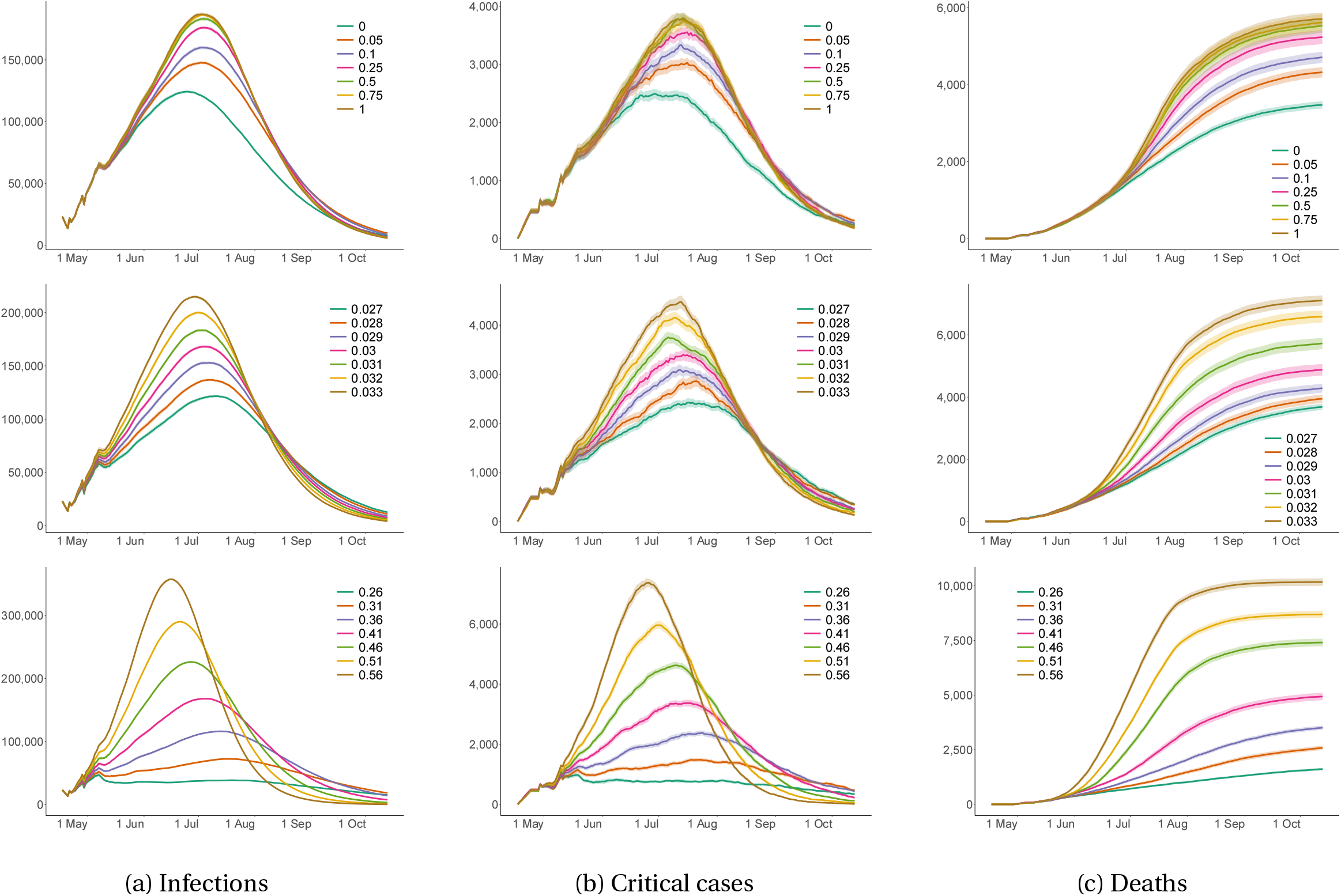
Key observables as a function of varying key parameters. This figure show infections, critical cases and deaths for Cape Town in a Lockdown scenario, for different values of the weight, *ρ*, of the private signal, *ζ*_*j*_ (*t*), in the agent’s compliance decision (top), the probability of transmission, *π*^*E*^ (middle) and the minimum share of contacts made under the most stringent lockdown, *ω*. All figures show the mean outcome taken across 50 simulations. For reference, in our baseline specifications, *ρ* = 0.15, *π*^*E*^ = 0.030 and *ω* = 0.46. The shaded area indicates the standard deviation.

When we compare two extreme cases: restricting the model to only social learning (*ρ* = 0) merely moves the date of peak infections forward by 2 days relative to a model without social learning (*ρ* = 1). While reducing the weight on the private signal reduces infections, there is a strong non-linearity in this relationship when the private signal is eliminated altogether (*ρ* = 0). In fact, going from *ρ* = 1 to *ρ* = 0.05 reduces total infections by 10%, while going from *ρ* = 0.05 to *ρ* = 0 reduces total infections by 16%.

On the other hand, reducing the transmission probability reduces the number of infections as well as the number of peak infections and also moves back the date of peak infections. When compared to a simulation with *π*^*E*^ = 0.33, a simulation with *π*^*E*^ = 0.27 produces 17% lower total infections, 43% lower peak infections and moves the date of peak infections back by 9 days. In addition, the virus also takes longer before it dies out completely. As a result, changes in the two uncertain parameters *ρ* and *π*^*E*^ produce quantitatively different effects, while both an an increase in *ρ* and a reduction in *π*^*E*^ reduce infections, the former brings forward the date of the peak while the latter pushes it back in time.

The fact that social learning reduces the height of the curve without stretching it out can be explained by the fact that agents will only increase their compliance, and hence reduce their contacts, once the virus has affected their neighbors in the social network. That being said, lockdown policies by the government are also a major factor that determines compliance and contacts. As Figure 5 shows, increasing the weight of social learning non-linearly increases compliance and reduces contacts, especially once the virus became widespread in July 2020. Finally, as can be seen in the figure, with a full weight on social learning the increased rate of fatalities results in almost full compliance by agents.

**Figure 5:**
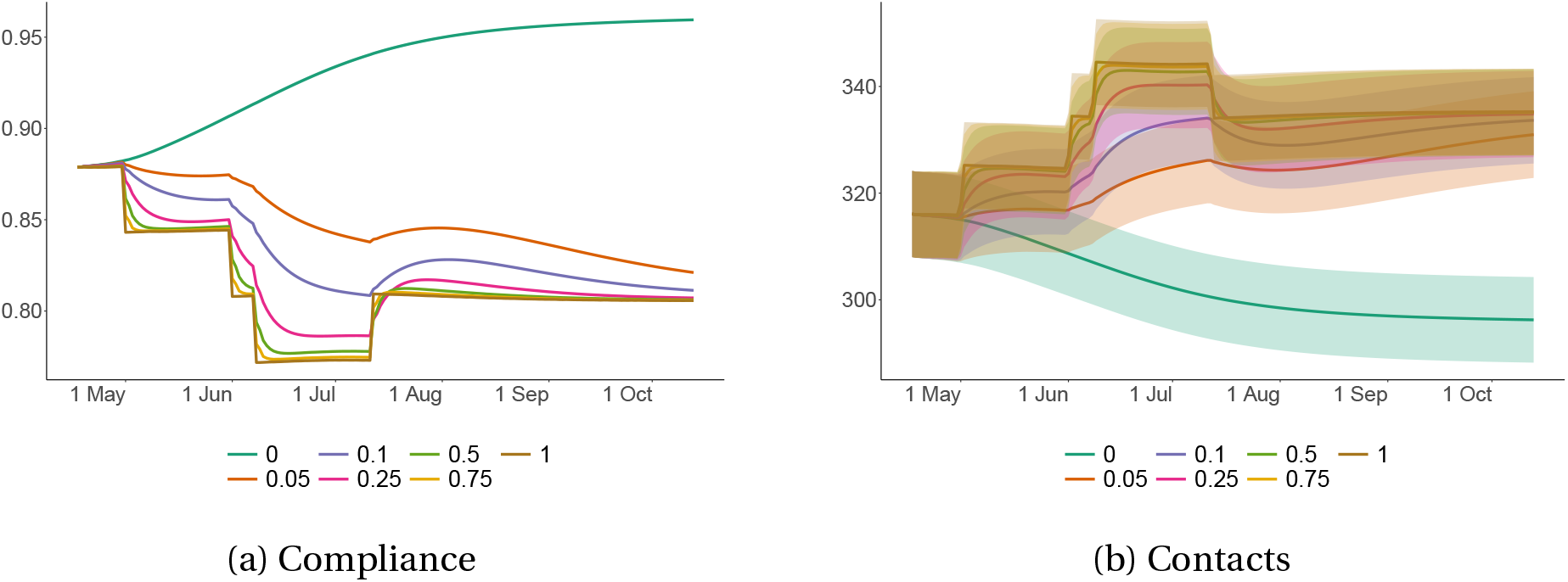
Compliance and number of contacts for varying relative strength of the private signal. This figure show compliance and contacts for Cape Town in a Lockdown scenario, for different values of the weight, *ρ*, of the private signal, *ζ*_*j*_ (*t*), in the agent’s compliance decision. All figures show the mean outcome taken across 50 simulations. For reference, in our baseline specifications, our calibrated value of *ρ* is 0.15. The shaded area indicates the standard deviation.

### 4.3 Social Learning and Government Intervention

Finally, since our calibration period coincided with government lockdown policies, we examine how social learning affects the spread of the virus in the–hypothetical–absence of such measures.

We consider two scenarios. The first scenario is our baseline, the scenario discussed in Section 3. We contrast this with a second *‘no-intervention’* scenario in which we assume no interventions by the government, meaning that the private signal *ξ*(*t*) = 0 for *t* ∈ *T*. This allows us to both analyze the effectiveness of the government lockdown, which is important to quantify, given the high economic cost the lockdown had in South Africa (Arndt et al., 2020), as well as the effect of social learning when the government does not intervene.

First, we consider the effect of the lockdown in the model with social learning. In the no-intervention scenario, infections among the population exceed 3 million, with 81% of the population becoming infected. Peak infections are reached on day 43 of the simulation and the healthcare system becomes overwhelmed, with the number critical cases exceeding the hospital capacity for 51 days, resulting in additional deaths as a result of insufficient healthcare. By the end of the pandemic, there are 12,973 deaths resulting in a infection fatality ratio (IFR) of 0.43%. Panel (e) in Figure 6 shows the evolution of compliance. Compliance takes a similar shape to infections and critical cases and compliance approaches it’s peak roughly at the same time as infections and agents begin to reduce the number of contacts they make, as illustrated in Panel (f). This endogenous increase in compliance and a reduction in contacts in response to rising infections illustrates the effect of social learning in our model. Despite this, social learning happens too slowly and always lags infections—once compliance reaches it’s peak, infections have already reached their peak. This highlights an important role for policy in the form of a lockdown: increasing compliance when infections are low, in anticipation of a rise in infections in the near future.

**Figure 6:**
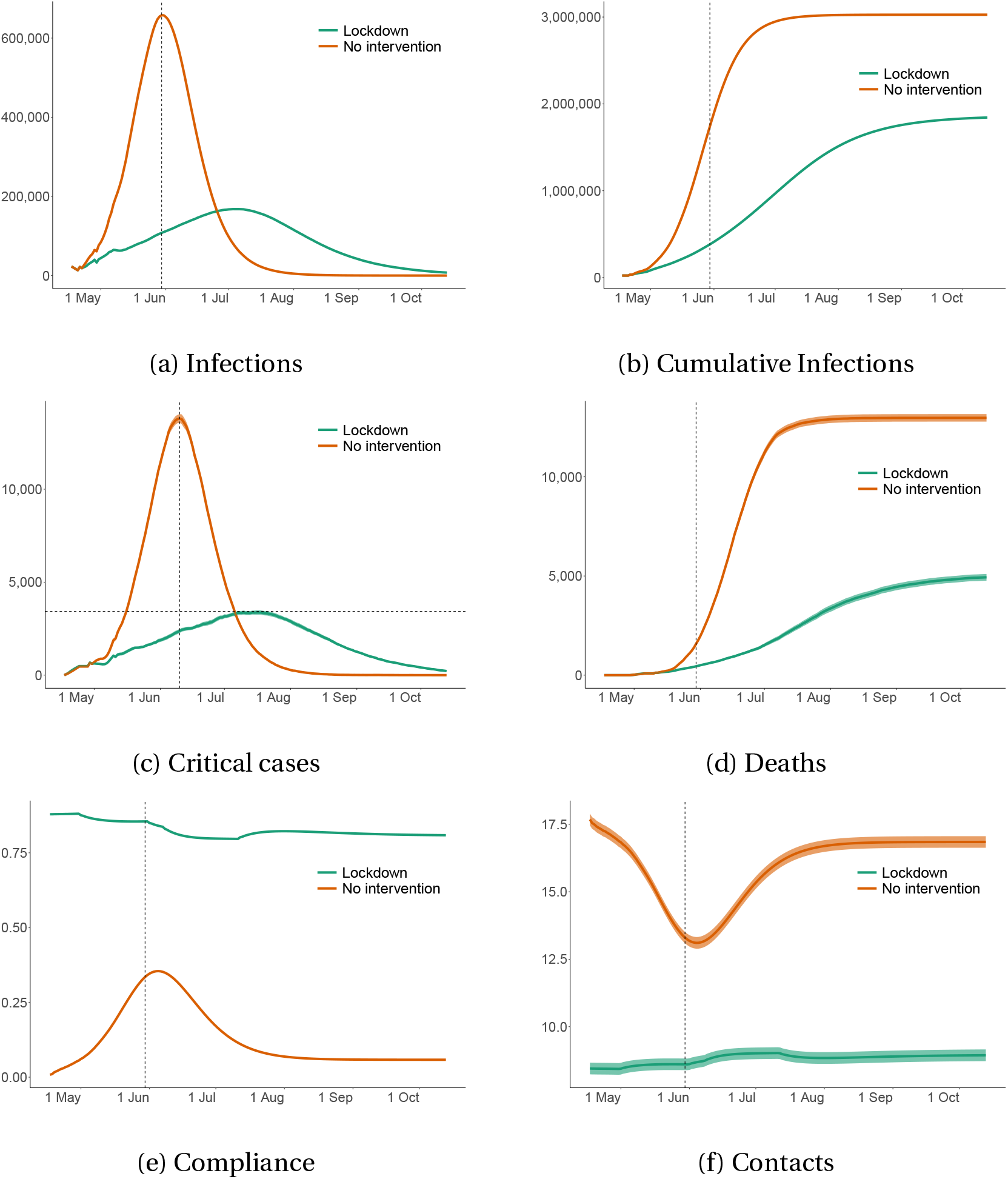
The impact of a lockdown on key model observables. This figure shows simulation outcomes for Cape Town across the lockdown and no-intervention scenario. Panels (a)–(d) show the number of infections, cumulative infections, critical cases, and deaths, respectively. Numbers are scaled to the size of Cape Town. Panel (e) shows the level of compliance and panel (f) shows the average number of contacts per household. Panels (a), (b), (c), (e), and (f) include vertical lines indicating the date of peak infections. In Panel (c) we also include a dotted horizontal line indicating hospital capacity in Cape Town. All figures show the mean outcomes taken across 50 simulations.

In the lockdown scenario, infections are significantly lower, with infections reaching 1.8 million, resulting in 49% of the population becoming infected. The trajectory of infections is also quantitatively different from the trajectory in a no-intervention scenario: the curve is flatter (i.e. smaller peak and larger standard deviation) and the virus takes longer to die out completely. Peak infections are reached on day 79, more than five weeks later than in a no-intervention scenario. Furthermore, peak infections and peak critical cases are significantly lower, ensuring that the healthcare system is only overburdened for 12 days. Altogether, this results in 4,934 deaths and an IFR of 0.27%, around four times lower than the IFR in a no-intervention scenario. Compliance behaviour is also distinctly different under a lockdown, as illustrated in Panel (e). At the beginning of the simulation, compliance is already high, taking a value of around 0.8, compared to a mechanical value of 0 in the no-intervention scenario. This is due to the fact that, under a lockdown, compliance becomes a function of a private signal, *ζ*_*j*_ (*t*) which reflects lockdown stringency and social learnin 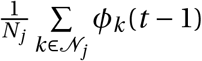, whereas in the no-intervention scenario, the private signal equals zero by definition, *ζ*_*j*_ (*t*) = 0. As a result, the introduction of a lockdown leads to an upward shift in compliance early in the pandemic, at precisely the time where infections and compliance due to social learning is low. Compliance remains high throughout the pandemic as the lockdown (with varying intensity at different times) remains intact.

These results highlight that four important features of a lockdown are still present in a network model with social learning. First, lockdown regulations reduce mobility and thus infections and consequently the number of deaths. Second, they have an additional dampening effect on deaths by limiting the extent to which the hospital system becomes overburdened, reducing the number of preventable deaths which arise solely due to inadequate access to health services. Third, lockdown regulations delay the peak of the pandemic, providing additional time for policymakers to prepare for the pandemic. One benefit of this additional time is that it gives policymakers time to expand the capacity of the healthcare system. Finally, lockdowns increase compliance early in the pandemic when endogenous compliance due to social learning is low. While social learning is effective inducing endogenous increases in compliance, independent of policy action, it always happens at a lag to infections, reducing its effectiveness in suppressing infections. Lockdowns create the initial increase in compliance and a reduction in contacts necessary to suppress the extent to which the virus seeds. Highlighting this effect is a key contribution of our paper.

Next, we test how varying the weight of social learning affects the virus curves in the no-intervention scenario and compare it to our baseline. Comparing Figure 7 to Figure 4, the same basic patterns emerge when we vary the weight of the private signal *ρ*. Introducing and increasing the extent of social learning lowers the peak of the infections, but does not change the duration of the pandemic. For example, comparing the two extreme cases, a model with only social learning (*ρ* = 0) merely moves the date of peak infections forward by 2 days relative to a model without social learning (*ρ* = 1). While reducing the weight on the private signal reduces infections, there is a strong non-linearity in this relationship when the private signal is eliminated altogether (*ρ* = 0). In fact, going from *ρ* = 1 to *ρ* = 0.05 reduces total infections by 10%, while going from *ρ* = 0.05 to *ρ* = 0 reduces total infections by 16%. However, the same *ρ* = 0.05 to *ρ* = 0 reduction only results in a 7.3% decrease in total infections in the no-intervention scenario.

**Figure 7:**
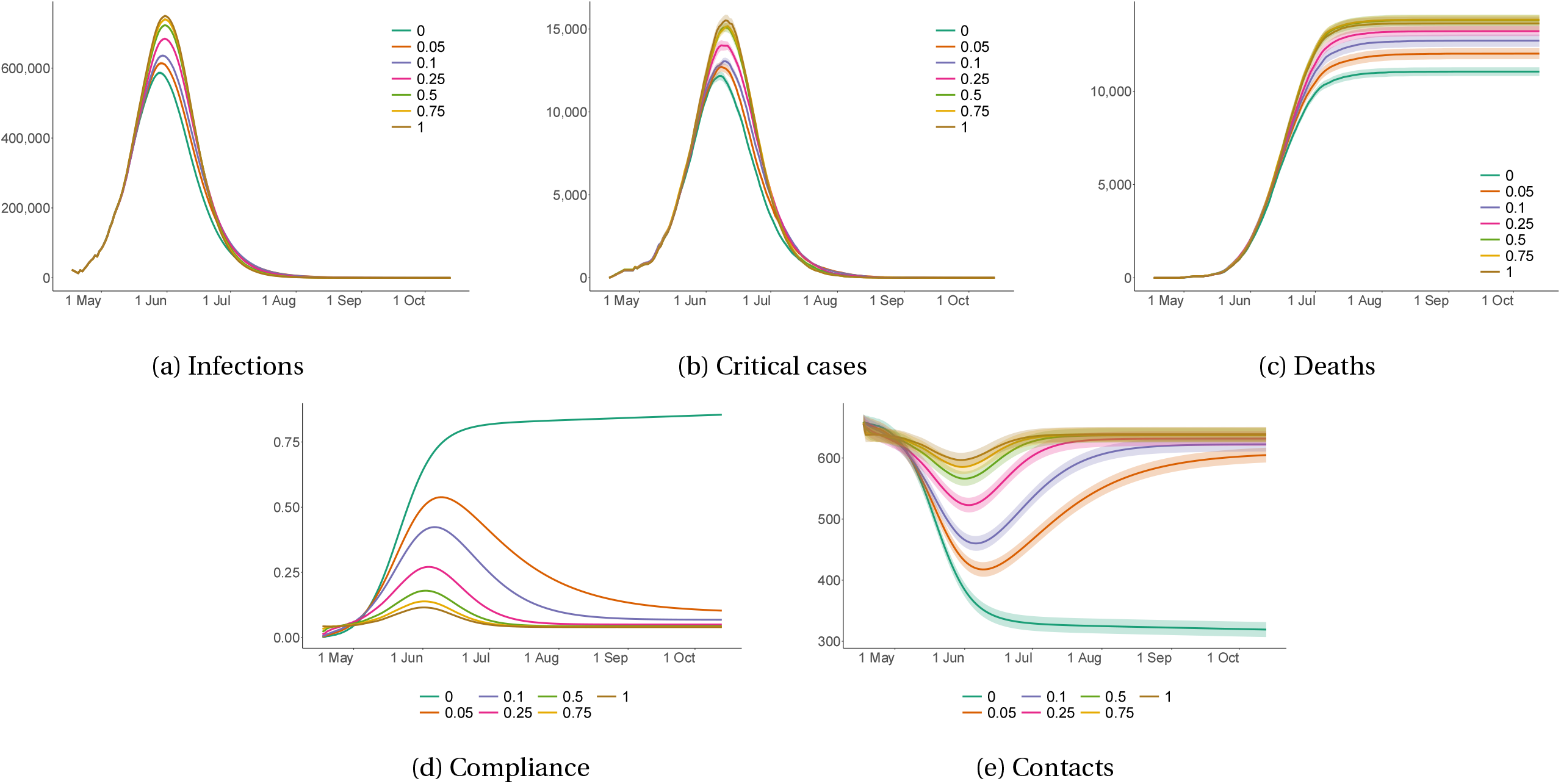
Key observables as a function of a changing weight of the private signal. This figure show infections, critical cases, deaths compliance and contacts for Cape Town in a no-intervention scenario, for different values of the weight, *ρ*, of the private signal, *ζ*_*j*_ (*t*), in the agent’s compliance decision. All figures show the mean outcome taken across 50 simulations. Shaded areas indicate the standard deviation. For reference, in our baseline specifications, our calibrated value of *ρ* is 0.15

The biggest difference is that now the private signal *ρ* = 0 acts as an anchor for compliance. Again, compliance will remain higher only in the extreme case of full weight on social learning. In all other cases compliance tends to go back to zero once the virus subsides. As a result, increasing the weight of the social signal mechanically increases compliance as indicated in Panel (c) of Figure 7. However, when comparing *ρ* = 0.05 to *ρ* = 0, the difference in compliance as the virus takes off all the way through to the peak of infections is small, with the major differences occurring after the peak of the virus. It also worth discussing the two polar cases, namely *ρ* = 0 and *ρ* = 1. When *ρ* = 0, agents base their compliance solely on social learning. Compliance reaches a peak after infections peak and remains at this peak. Given the fact that in a no-intervention scenario, deaths are significantly higher, agents are much more likely to be exposed to a death in their network of connections and since we encode the compliance of dead agents to be 1 at all times after their death, this causes aggregate compliance to converge to a high level even as the pandemic ends. Contrast this with the case where *ρ* = 0. In this scenario, by default all agents do not comply with the lockdown policies and do not reduce their contacts. Agents only increase their compliance, when they become infected. If agents die, their compliance remains 1 throughout, while recovered agents revert to the non-compliance norm. As a result, compliance is always non-zero due to initial infections and deaths, and there is a slight increase in compliance driven by increased compliance among the infected population around the peak of the pandemic.

Now, contrast the compliance behaviour in the lockdown scenario. In all cases, compliance already begins at a high level, driven the stringency of the lockdown which affects the private signal. In the special case where *ρ* = 1, compliance simply becomes the private signal, which is calibrated to the lockdown stringency index. This explains the step changes in compliance behaviour. As soon as social learning is introduced (*ρ* < 1), compliance is smoothed. In the case of social learning only, *ρ* = 0, compliance once again plateau’s at a high level, with the transition to this plateau being more gradual than the no intervention case, driven by the slower progression of infections. However, unlike the no-intervention scenario, here there are clear differences in compliance early in the pandemic when comparing *ρ* = 0.05 to *ρ* = 0. Put differently, under a lockdown, when agents only make use of social learning this leads to a significant increase in compliance, even when compared to a calibration where a small weight is placed on the private signal. This change in compliance happens early enough to meaningfully impact infections, explaining the larger decrease in total infections (16%) when compared to the no-intervention scenario (7.3%).

The main insight from these results is that the inclusion of social learning has significant effects on the projected progression of the virus. Comparing results from simulations using our calibrated specification, *ρ* = 0.15 with an identical model without social learning *ρ* = 1, leads to a 4% decrease in predicted infections and a 17.4% decrease in predicted deaths. The more pronounced reduction in deaths stems from the fact that in the model with social learning, critical cases never exceed healthcare capacity whereas in the model without social learning, the healthcare system is overburdened for 28 days, resulting in many preventable deaths.

## 5 The Effectiveness of Different Vaccination Strategies

Covid-19 network models are particularly useful to study the efficacy of different vaccination strategies because there is no heterogeneous mixing and therefore local herd immunity can be achieved. Globally, the most common vaccination strategy (World Health Organization and others, 2020) is risk-based. Under such a strategy, particularly vulnerable agents—for example the elderly or those with comorbidities—are first in line to be vaccinated after healthcare and front-line workers. The rationale behind this strategy is that a relatively small part of the population is particularly susceptible to being severely affected by Covid-19. By vaccinating those first, governments can quickly prevent hospitals from being overwhelmed and effectively reduce the death toll. One possible alternative, put forward by Bubar et al. (2021), is to protect those most at risk indirectly by vaccinating those who transmit the virus most widely. The idea is that the spread of the disease can be effectively curbed if those who are most likely to transmit it are vaccinated.^38^ Using our calibrated model, we compare the efficacy of these two vaccination strategies with a third vaccination strategy wherethe order in which age categories are vaccinated is random. Specifically, we study how these three vaccination strategies affect fatalities, critical cases, and infections.

We explore a hypothetical scenario in which one-shot perfectly effective vaccines are available at the start of our simulation. This means that each agent who is vaccinated will immediately change her status to recovered. For simplicity, we assume that only susceptible agents will be vaccinated. Furthermore, we assume that, while the vaccine technology is available at the start of the simulation, only 500 vaccines can be produced and distributed each period (i.e. per day). This implies that the 100,000 agents in our simulation are vaccinated within 200 days.

The shortage of vaccines implies that a choice needs to be made about which agents get vaccinated first. For our experiments, we first divide agents into their respective age classes: 0 − 10, 10−20,…, 80+. For the risk-based strategy, agents within the age class are vaccinated in random order. For the connection-based strategy, we use the contact matrix from Table 2, which captures the probability that an agent in a particular age group interacts with an agent from another age group. We use this information as a proxy for an agent’s connectedness. Lastly, for the random vaccination strategy, each day 500 agents are selected at random, irrespective of age.

For each of these vaccination scenarios, we simulate our calibrated model with *V* = 50 Monte Carlo simulations, shown in Figure 8. We find that, while the vaccination speed is too slow to completely eliminate the virus, the curves are flattened considerably by all three. When comparing the three strategies, the connection-based strategy is the most effective at controlling infections. If this effect was very strong, it could lead to a large reduction in total fatalities, possibly outweighing the reduction in fatalities under the risk-based strategy.

**Figure 8:**
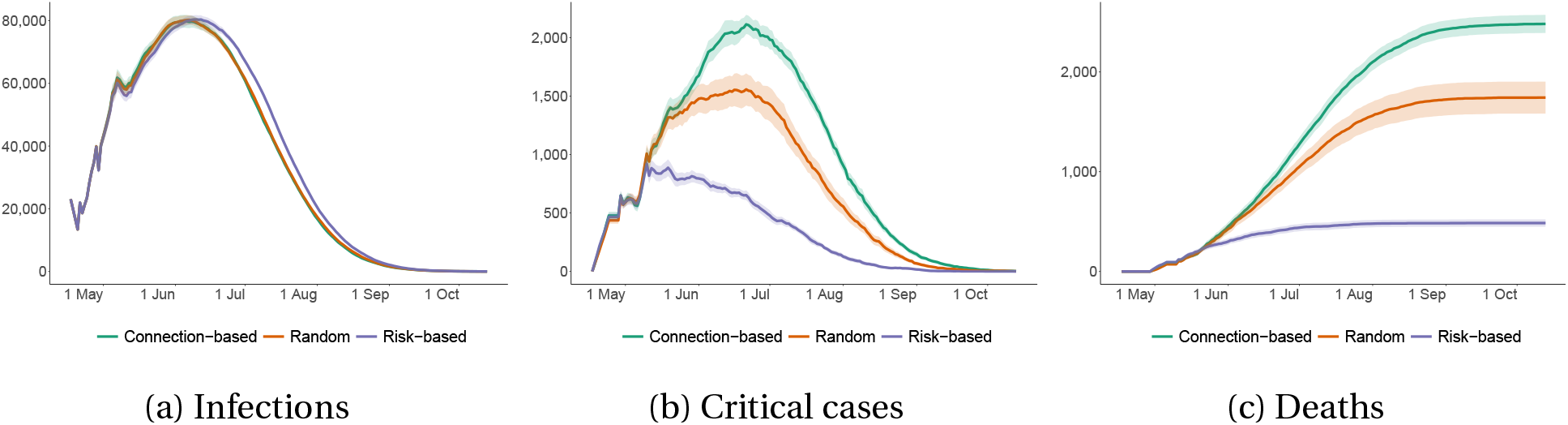
The efficacy of different vaccination strategies. This figure shows infections, critical and deaths agents over time, under the three vaccination scenarios: connection based; random and risk-based. All figures show mean results across 50 simulations. The shaded regions represent 95% confidence bands.

This, however, is not the case, on average infections are hardly influenced by the different strategies. As shown in Table 8, the connection-based strategy leads to 0.9% less infections than the random strategy, while the risk-based strategy leads on average, leads to a 5.3% *increase* in cumulative infections. Even when to compared to the risk-based strategy, the connection based strategy barely leads to a 5.9% decrease in infections.

On the other hand, there are large differences in fatalities. Compared to the random benchmark, the connection-based vaccination strategy results in 42% *more* fatalities than the random vaccination strategy, while the risk-based scenario leads to a reduction of roughly 72%. Comparing the two scenarios leads to an even more stark difference. By changing from a connection-based to a risk-based vaccination strategy, fatalities can be reduced by a little over 80%.

Since both total infections and the government stringency hardly changes in these scenarios, the differences in compliance and contacts are very small.

## 6 Conclusion

Our simulations show that social learning helps explain the unusually flat and short Covid-19 curves in Cape Town. The social learning signal is strong when there are many infections in the neighbourhood of an agent. This decreases the spread of Covid-19 at the peak and since agents comply for a while, social learning speeds up the decline of the virus. This is different from the effect of lockdown measures that reduce the transmission probability, which reduce total infections by spreading them out over time. Nevertheless, social learning alone was not enough to flatten the curve in Cape Town so that hospital capacity was not breached, increasing the excess fatality rate. A lockdown was still needed to flatten the curve sufficiently. However, the reduction in the number of contacts as a consequence of a lockdown is smaller when taking into account social learning because otherwise agents will also reduce some contacts voluntarily. Finally, we show that in our calibrated model the strategy of vaccinating the elderly first effectively reduces the number of fatalities, even though there are more infections compared to a random strategy, and even more so compared to a strategy in which the most connected agents are vaccinated first.

This paper has three important insight for policy makers. First, standard epidemiological models without social learning are biased to overstate either the height or length of the pandemic. As a consequence, these models overestimate the loss of life due to the pandemic because they do not consider that in the no-intervention scenario people will also voluntarily reduce their contacts as a result of social learning. Second, models without social learning overestimate the cost of lockdown measure in terms of reduced contacts. Finally, a risk-based vaccination strategy is highly effective and leads to much lower fatalities, even in cities with relatively young populations such as Cape Town.

Having been fully calibrated, the model presented in this paper offers many possibilities for extensions which will make it applicable to particular policy concerns. For example, agent behaviour can easily be modified to include some sort of trade-off between contacts and age specific risks. Furthermore, thanks to its detailed heterogeneity, the model lends itself well to policy experiments that target specific areas or age profiles.

## Data Availability

All data and scripts are available on Github

https://github.com/blackrhinoabm/sabcom

## Appendix A Tables

**Table 3:** Simulation results under both the lockdown and no-intervention scenario We include (i) Total Infections (ii) Total Deaths (iii) Total Recoveries (iv) Peak Infections (v) Peak Critical (vi) Peak Infections (Day) - the day on which peak infections is reached (vii) Peak Critical (Day) - the day on which peak critical is reached, (viii) End of simulation infections - the number of agents still infected at the end of the simulation, (ix) Critical > Capacity (Days) - the number of days in which the number of critical cases exceeds hospital capacity, (x) the average number of contacts and (xi) the average level of compliance. We report minimum, mean and maximum values, as well as the standard deviation across 50 simulations.

| Outcome | Min | Mean | Max | SD | Min | Mean | Max | SD |
| --- | --- | --- | --- | --- | --- | --- | --- | --- |
| Total Infections | 1,801,982 | 1,843,002 | 1,889,125 | 22,008 | 3,010,721 | 3,026,825 | 3,041,277 | 7,938 |
| Total Deaths | 3,740 | 4,934 | 6,582 | 591 | 11,818 | 12,973 | 14,736 | 665 |
| Total Recoveries | 1,788,293 | 1,830,547 | 1,879,363 | 22,474 | 2,997,893 | 3,013,849 | 3,028,187 | 7,782 |
| Peak Infections | 155,735 | 167,847 | 176,941 | 4,816 | 639,881 | 657,681 | 675,149 | 6,661 |
| Peak Critical | 2,655 | 3,368 | 4,413 | 409 | 11,744 | 13,803 | 16,344 | 827 |
| Peak Infections (Day) | 67 | 79 | 87 | 4 | 42 | 43 | 46 | 1 |
| Peak Critical (Day) | 54 | 86 | 105 | 10 | 48 | 54 | 57 | 2 |
| End of simulation infections | 5,124 | 7,520 | 10,098 | 1,243 | 0 | 4 | 37 | 11 |
| Critical > Capacity (Days) | 0 | 12 | 30 | 9 | 47 | 51 | 54 | 1 |
| Contacts | 7.3 | 8.87 | 10.61 | 0.76 | 14.76 | 16.31 | 18.06 | 0.76 |
| Compliance | 0.82 | 0.82 | 0.82 | 0 | 0.1 | 0.1 | 0.1 | 0 |
| (a) Lockdown |  |  |  |  | (b) No-intervention |  |  |  |

**Table 4:** Simulation results when varying the extent of social learning in a lockdown scenario This table shows model outcomes for Cape Town for different values of the weight, *ρ*, of the private signal, *ζ*_*j*_ (*t*), in the agent’s compliance decision in a lockdown scenario. We include (i) Total Infections (ii) Total Deaths (iii) Total Recoveries (iv) Peak Infections (v) Peak Critical (vi) Peak Infections (Day) - the day on which peak infections is reached (vii) Peak Critical (Day) - the day on which peak critical is reached, (viii) End of simulation infections - the number of agents still infected at the end of the simulation, (ix) Critical > Capacity (Days) - the number of days in which the number of critical cases exceeds hospital capacity, (x) the average number of contacts and (xi) the average level of compliance. We report minimum, mean and maximum values, as well as the standard deviation across 50 simulations.

| Outcome | $\rho = 0$ | $\rho = 0.05$ | $\rho = 0.1$ | $\rho = 0.15$ | $\rho = 0.25$ | $\rho = 0.5$ | $\rho = 0.75$ | $\rho = 1.0$ |
| --- | --- | --- | --- | --- | --- | --- | --- | --- |
| Total Infections | 1,450,666 | 1,726,480 | 1,803,981 | 1,843,002 | 1,875,291 | 1,903,463 | 1,915,957 | 1,919,512 |
| Total Deaths | 3,477 | 4,326 | 4,714 | 4,934 | 5,236 | 5,534 | 5,623 | 5,710 |
| Total Recoveries | 1,439,665 | 1,712,503 | 1,790,852 | 1,830,547 | 1,863,480 | 1,891,933 | 1,904,569 | 1,908,184 |
| Peak Infections | 124,312 | 147,684 | 159,934 | 167,847 | 176,092 | 183,248 | 186,619 | 186,885 |
| Peak Critical | 2,495 | 3,028 | 3,331 | 3,368 | 3,555 | 3,800 | 3,749 | 3,778 |
| Peak Infections (Day) | 69 | 76 | 77 | 77 | 78 | 77 | 77 | 77 |
| Peak Critical (Day) | 73 | 90 | 87 | 90 | 90 | 88 | 88 | 85 |
| End of simulation infections | 7,523 | 9,651 | 8,415 | 7,520 | 6,575 | 5,996 | 5,765 | 5,618 |
| Critical > Capacity (Days) | 0 | 0 | 0 | 0 | 16 | 22 | 25 | 28 |
| Contacts | 8.02 | 8.75 | 8.83 | 8.87 | 8.9 | 8.92 | 8.93 | 8.93 |
| Compliance | 0.94 | 0.84 | 0.82 | 0.82 | 0.81 | 0.81 | 0.81 | 0.81 |

**Table 5:** Simulation results when varying the extent of social learning in a no-intervention scenario This table shows model outcomes for Cape Town for different values of the weight, *ρ*, of the private signal, *ζ*_*j*_ (*t*), in the agent’s compliance decision in a no-intervention scenario. We include (i) Total Infections (ii) Total Deaths (iii) Total Recoveries (iv) Peak Infections (v) Peak Critical (vi) Peak Infections (Day) - the day on which peak infections is reached (vii) Peak Critical (Day) - the day on which peak critical is reached, (viii) End of simulation infections - the number of agents still infected at the end of the simulation, (ix) Critical > Capacity (Days) - the number of days in which the number of critical cases exceeds hospital capacity, (x) the average number of contacts and (xi) the average level of compliance. We report minimum, mean and maximum values, as well as the standard deviation across 50 simulations.

| Outcome | $\rho = 0$ | $\rho = 0.05$ | $\rho = 0.1$ | $\rho = 0.15$ | $\rho = 0.25$ | $\rho = 0.5$ | $\rho = 0.75$ | $\rho = 1.0$ |
| --- | --- | --- | --- | --- | --- | --- | --- | --- |
| Total Infections | 2,650,236 | 2,858,741 | 2,964,293 | 3,026,770 | 3,094,962 | 3,166,939 | 3,194,733 | 3,209,114 |
| Total Deaths | 11,052 | 12,016 | 12,710 | 12,853 | 13,220 | 13,804 | 13,837 | 13,633 |
| Total Recoveries | 2,639,184 | 2,846,712 | 2,951,578 | 3,013,914 | 3,081,743 | 3,153,135 | 3,180,897 | 3,195,481 |
| Peak Infections | 586,456 | 613,925 | 635,849 | 657,669 | 684,372 | 723,142 | 739,804 | 749,504 |
| Peak Critical | 12,167 | 12,730 | 13,054 | 13,739 | 14,014 | 15,087 | 15,173 | 15,518 |
| Peak Infections (Day) | 42 | 42 | 43 | 44 | 44 | 44 | 44 | 44 |
| Peak Critical (Day) | 52 | 52 | 54 | 54 | 52 | 54 | 55 | 54 |
| End of simulation infections | 0 | 13 | 4 | 3 | 0 | 0 | 0 | 0 |
| Critical > Capacity (Days) | 47 | 49 | 51 | 50 | 50 | 49 | 50 | 49 |
| Contacts | 10.34 | 14.56 | 15.49 | 15.89 | 16.27 | 16.67 | 16.8 | 16.87 |
| Compliance | 0.67 | 0.24 | 0.16 | 0.13 | 0.1 | 0.07 | 0.06 | 0.05 |

**Table 6:** Simulation results when varying the transmission probability in a lockdown scenario This table shows model outcomes for Cape Town for different values of the disease probability of transmission, *π*^*E*^ in a lockdown scenario. We include (i) Total Infections (ii) Total Deaths (iii) Total Recoveries (iv) Peak Infections (v) Peak Critical (vi) Peak Infections (Day) - the day on which peak infections is reached (vii) Peak Critical (Day) - the day on which peak critical is reached, (viii) End of simulation infections - the number of agents still infected at the end of the simulation, (ix) Critical > Capacity (Days) - the number of days in which the number of critical cases exceeds hospital capacity, (x) the average number of contacts and (xi) the average level of compliance. We report minimum, mean and maximum values, as well as the standard deviation across 50 simulations.

| Outcome | $\pi^E = 0.027$ | $\pi^E = 0.028$ | $\pi^E = 0.029$ | $\pi^E = 0.030$ | $\pi^E = 0.031$ | $\pi^E = 0.032$ | $\pi^E = 0.033$ |
| --- | --- | --- | --- | --- | --- | --- | --- |
| Total Infections | 1,528,530 | 1,642,133 | 1,745,812 | 1,843,391 | 1,929,874 | 2,018,460 | 2,095,866 |
| Total Deaths | 3,684 | 3,944 | 4,285 | 4,876 | 5,727 | 6,587 | 7,104 |
| Total Recoveries | 1,512,348 | 1,627,490 | 1,732,720 | 1,831,122 | 1,918,086 | 2,006,725 | 2,084,686 |
| Peak Infections | 121,602 | 136,931 | 152,983 | 168,051 | 183,468 | 200,145 | 214,966 |
| Peak Critical | 2,423 | 2,858 | 3,090 | 3,394 | 3,751 | 4,155 | 4,470 |
| Peak Infections (Day) | 85 | 81 | 81 | 78 | 77 | 75 | 73 |
| Peak Critical (Day) | 91 | 95 | 86 | 88 | 80 | 84 | 87 |
| End of simulation infections | 12,498 | 10,699 | 8,808 | 7,393 | 6,062 | 5,148 | 4,076 |
| Critical > Capacity (Days) | 0 | 0 | 0 | 0 | 23 | 36 | 44 |
| Contacts | 8.87 | 8.87 | 8.87 | 8.87 | 8.86 | 8.86 | 8.86 |
| Compliance | 0.82 | 0.82 | 0.82 | 0.82 | 0.82 | 0.82 | 0.82 |

**Table 7:** Simulation results when varying the transmission probability a no-intervention scenario This table shows model outcomes for Cape Town for different values of the disease probability of transmission, *π*^*E*^ in a no-intervention scenario. We include (i) Total Infections (ii) Total Deaths (iii) Total Recoveries (iv) Peak Infections (v) Peak Critical (vi) Peak Infections (Day) - the day on which peak infections is reached (vii) Peak Critical (Day) - the day on which peak critical is reached, (viii) End of simulation infections - the number of agents still infected at the end of the simulation, (ix) Critical > Capacity (Days) - the number of days in which the number of critical cases exceeds hospital capacity, (x) the average number of contacts and (xi) the average level of compliance. We report minimum, mean and maximum values, as well as the standard deviation across 50 simulations.

| Outcome | $\pi^E = 0.027$ | $\pi^E = 0.028$ | $\pi^E = 0.029$ | $\pi^E = 0.030$ | $\pi^E = 0.031$ | $\pi^E = 0.032$ | $\pi^E = 0.033$ |
| --- | --- | --- | --- | --- | --- | --- | --- |
| Total Infections | 2,894,028 | 2,941,954 | 2,985,157 | 3,028,015 | 3,064,321 | 3,102,929 | 3,133,917 |
| Total Deaths | 12,092 | 12,441 | 12,652 | 12,750 | 12,915 | 13,217 | 13,419 |
| Total Recoveries | 2,881,907 | 2,929,512 | 2,972,501 | 3,015,252 | 3,051,406 | 3,089,712 | 3,120,497 |
| Peak Infections | 574,730 | 603,624 | 630,594 | 657,482 | 682,598 | 709,875 | 734,193 |
| Peak Critical | 11,700 | 12,448 | 13,090 | 13,834 | 13,905 | 14,616 | 15,081 |
| Peak Infections (Day) | 46 | 45 | 44 | 44 | 43 | 42 | 42 |
| Peak Critical (Day) | 57 | 54 | 55 | 54 | 53 | 53 | 52 |
| End of simulation infections | 28 | 1 | 4 | 12 | 0 | 0 | 1 |
| Critical > Capacity (Days) | 52 | 51 | 51 | 50 | 50 | 49 | 50 |
| Contacts | 15.97 | 15.96 | 15.95 | 15.95 | 15.95 | 15.95 | 15.94 |
| Compliance | 0.12 | 0.13 | 0.13 | 0.13 | 0.13 | 0.13 | 0.13 |

**Table 8:** Vaccination experiments This table shows model outcomes for Cape Town under different vaccination scenarios in a lockdown scenario. We include (i) Total Infections (ii) Total Critical Cases (iii) Total Deaths (iv) Total Recoveries (v) Contacts and (vi) Compliance. We report mean values across 50 simulations.

| Outcome | Connection-based | Random | Risk-based |
| --- | --- | --- | --- |
| Total Infections | 902,706 | 911,008 | 959,736 |
| Total Critical cases | 545 | 413 | 201 |
| Total Deaths | 2,481 | 1,743 | 485 |
| Total Recoveries | 3,470,921 | 3,482,767 | 3,504,965 |
| Contacts | 8.9 | 8.9 | 8.9 |
| Compliance | 0.81 | 0.81 | 0.81 |

## Appendix B Pseudo Code: Initialisation Algorithm

Our algorithms can be split up into two main algorithms. First, we use initialisation Algorithm 1 that consists of three sub-algorithms.

### Algorithm 1 Initialisation

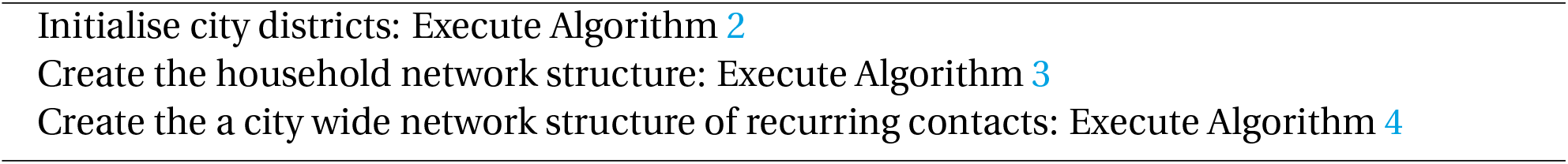

Next, we describe each of these sub-algorithms in more detail.

### B.1 Initialise City Districts

We use Algorithm 2 to calculate how many agents there should be in the simulation and what properties they should have to be proportional to the modelled city.

#### Algorithm 2 Initialise city districts

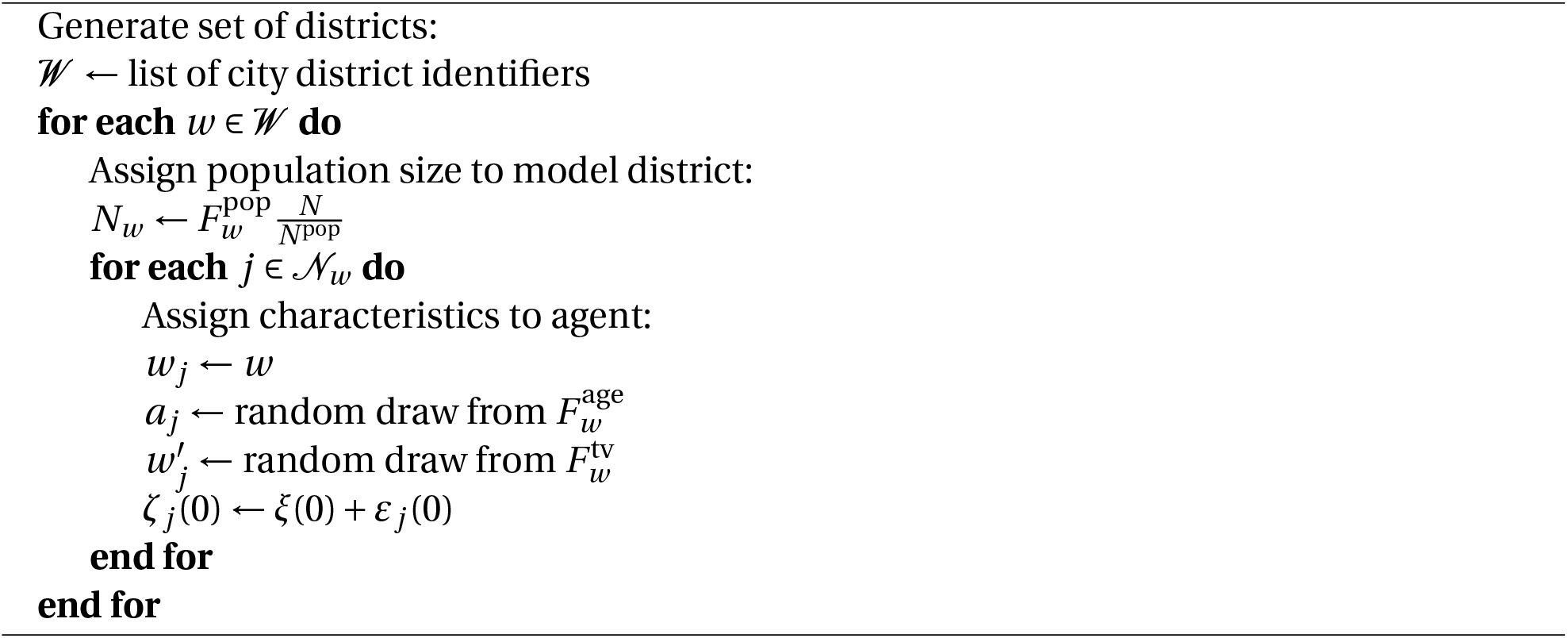

### B.2 Create the Household Network Structure

Next, the creation of the household network structure is described in Algorithm 3.

### B.3 Create the City Wide Network Structure of Other Contacts

Finally, algorithm 4 creates a city wide network for all non-household contacts. After these algorithms have been executed, we save the modelled city so it can be simulated.

## Appendix C Pseudo Code: Main Simulation Algorithm

An overview of the main simulation algorithm is described in algorithm 5

### C.1 Initial Infections

Algorithm 6 infects a number of initial agents. These agents are spread over the different districts in proportion to the initially detected cases per district from *F* ^ca^. All initially infected agents will have their initial status (*P* _*j*_ (0)) updated to exposed, infected without symptoms, or infected with symptoms, and the number of days for which they have been in the status at the start of the simulation 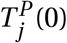 set as a random integer between zero and the maximum days that agents can be in that compartment.

#### Algorithm 3 Create the household network structure

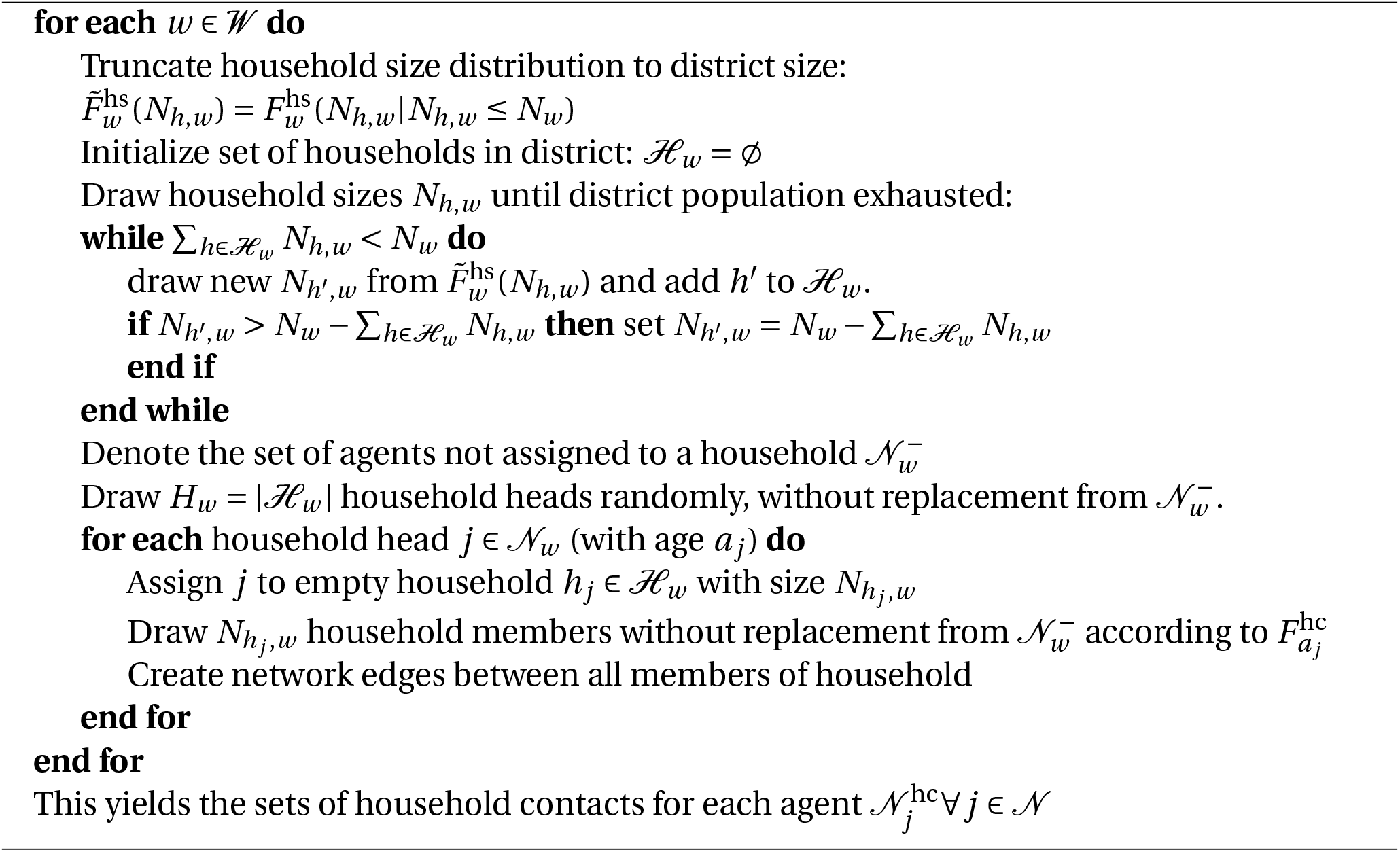

#### Algorithm 4 Create the other contacts network structure

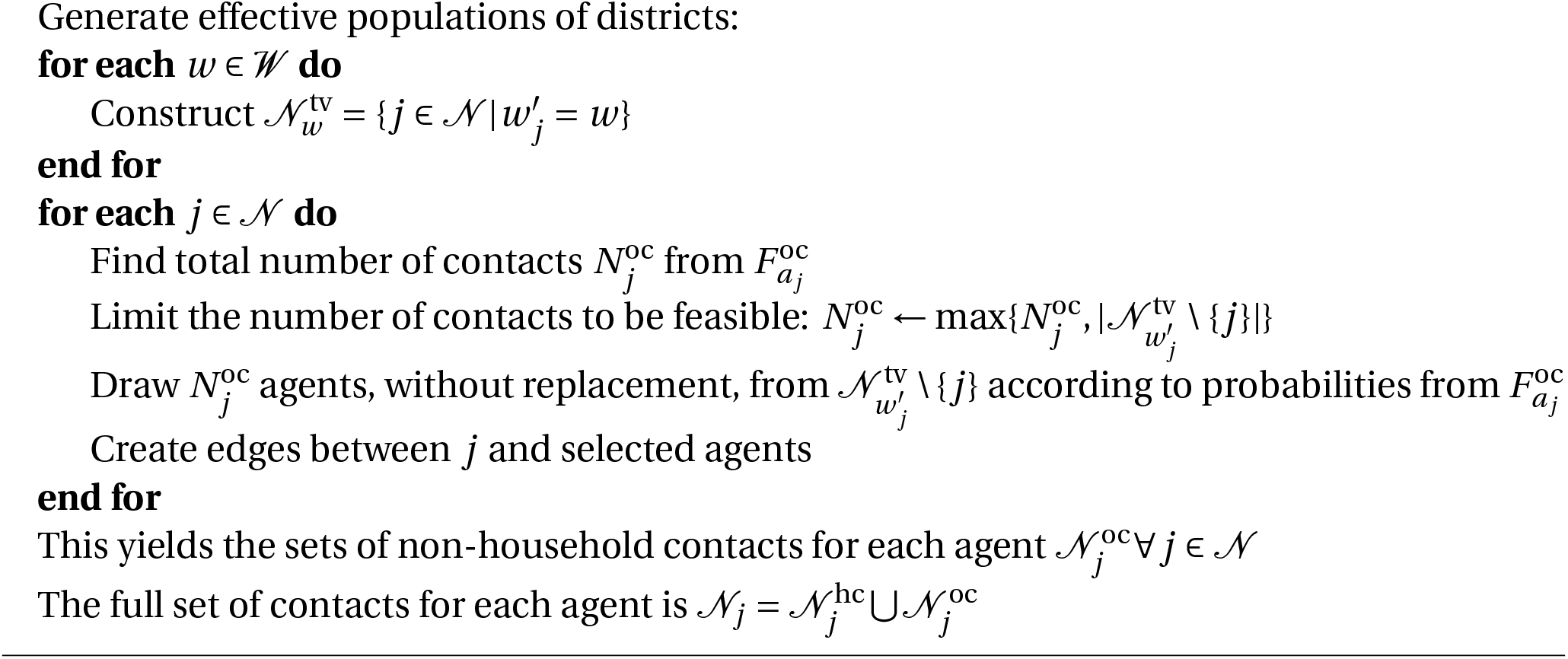

#### Algorithm 5 Main simulation

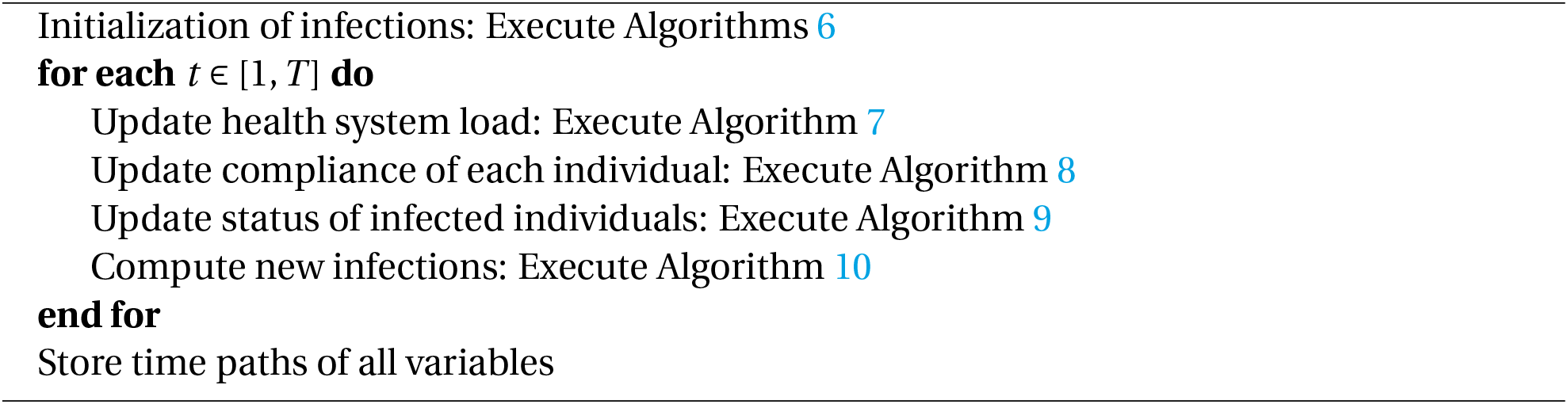

#### Algorithm 6 Initialize infections

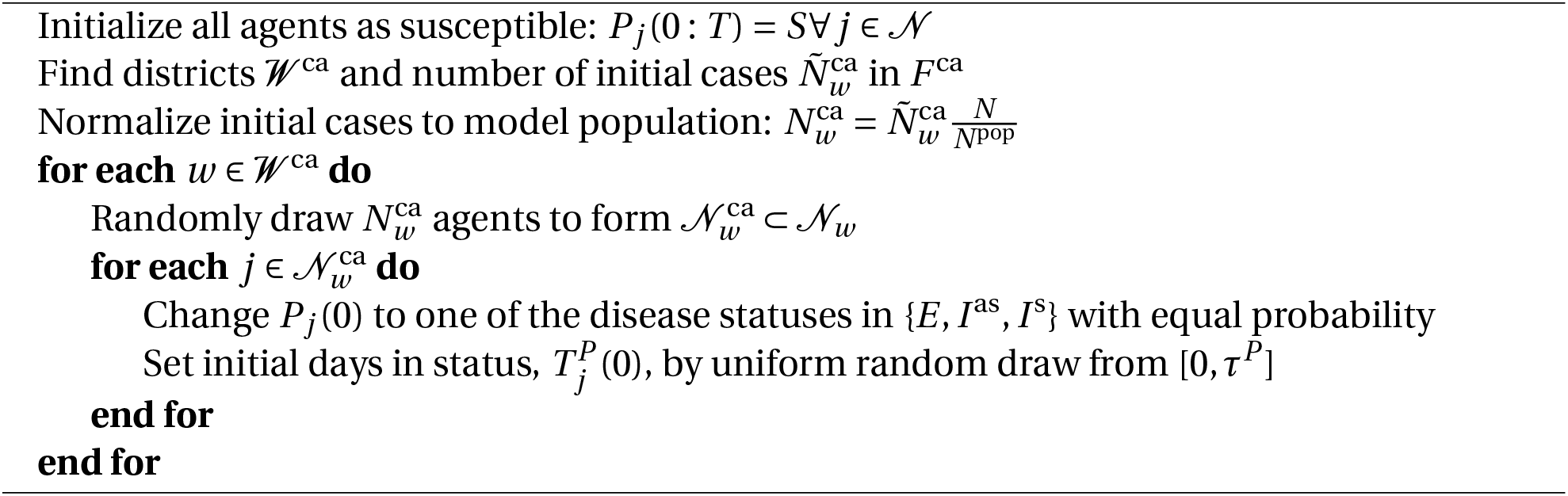

After the initial infections have occurred, the day loop simulation starts and the next algorithms will be called every day.

### C.2 Check Health System Capacity

Algorithm 8 checks if the health system is overburdened and activates the fatality multiplier *δ*^*L*^ if it is.

#### Algorithm 7 Check health system capacity

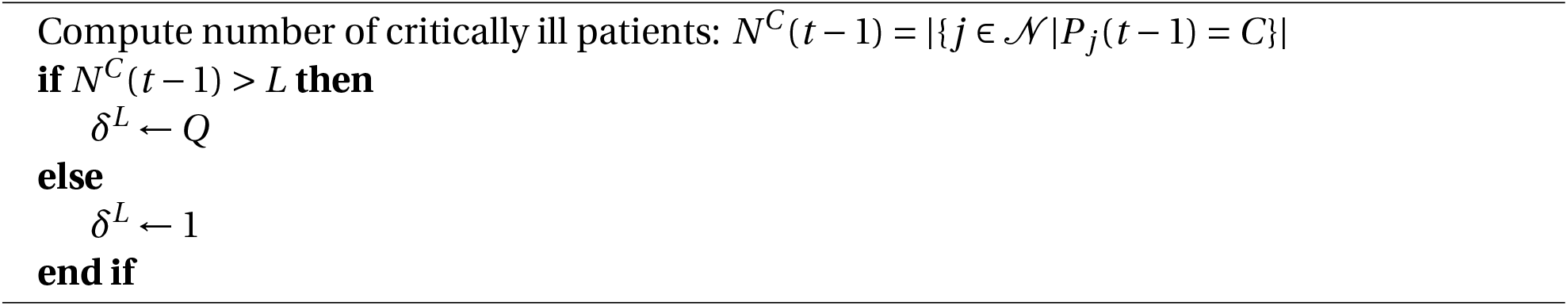

### C.3 Update Compliance

Next, every agent *j* updates its degree of compliance with lockdown regulations, through a naive deGroot learning Algorithm 8. Each agent has a private signal *ζ*_*j*_ (*t*) which consists of a public signal *ξ*(*t*) and an individual noise term *ε*_*j*_ (*t*), where the public signal is informed by the stringency index of the country being modelled *F* ^*strin*^(*t*). *ξ*(*t*) ∈ [0, 1] and *ε*_*j*_ (*t*) is drawn independently for each *j* from a truncated normal distribution with support [−*ξ*(*t*), 1 − *ξ*(*t*)] and mean 0 and variance *σ*, such that *ζ*_*j*_ (*t*) ∈ [0, 1].

The level of compliance *φ*_*j*_ (*t*) of the agents is a weighted average of the private signal *ζ*_*j*_ (*t*) (with weight of *ρ*) and the social signal (with weight of (1 − *ρ*)). The social signal is the simple average of observed previous (*t* − 1) compliance of all neighbours 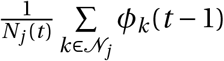.

#### Algorithm 8 Update agent compliance

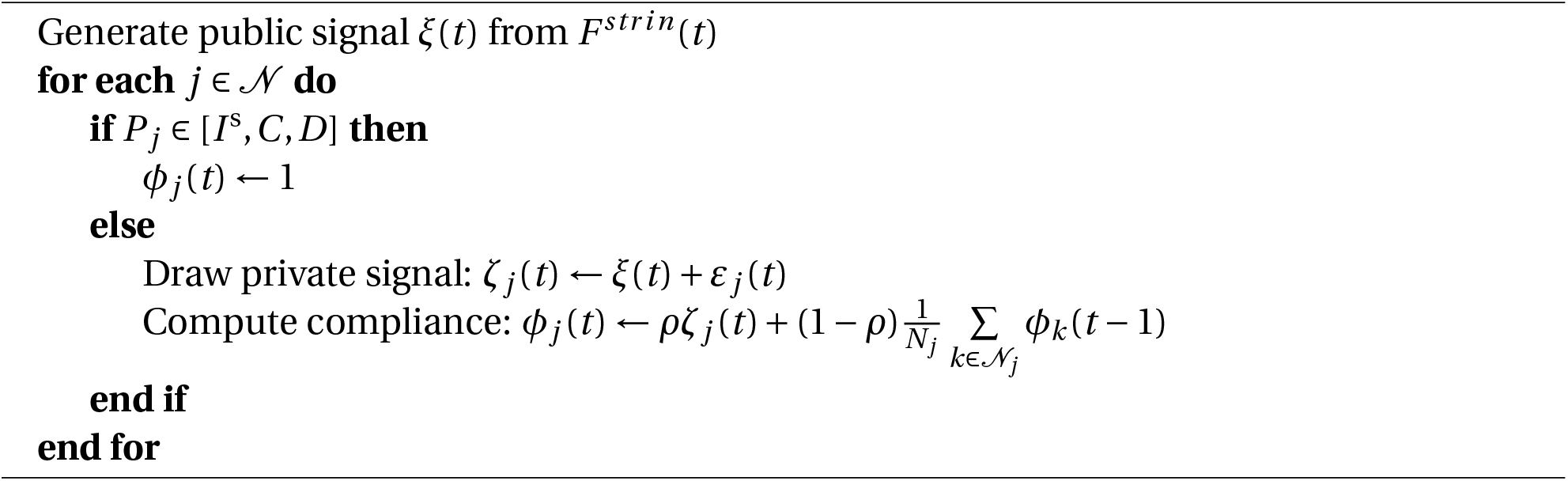

### C.4 Update Agents’ Infection Status

In Algorithm 9, each agent *j* that is in the exposed, asymptomatic, symptomatic, or critical compartments will update the number of days it has been in this status and, possibly, transition to a new disease status. Let 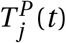 denote the number of days that agent *j* has been in disease compartment P by period *t* (where *τ*^*P*^ is the parameter that calibrates the tenure in compartment *P*). Since some transitions to different compartments are stochastic, we use random draws from a uniform distribution with support on [0, 1] to determine the outcome of a stochastic event *Z* for each agent in each period. We denote the random draw by 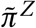. Thus, if the probability of event *Z* is *π*^*Z*^, then the event occurs only when 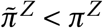.

#### Algorithm 9 Update infection status

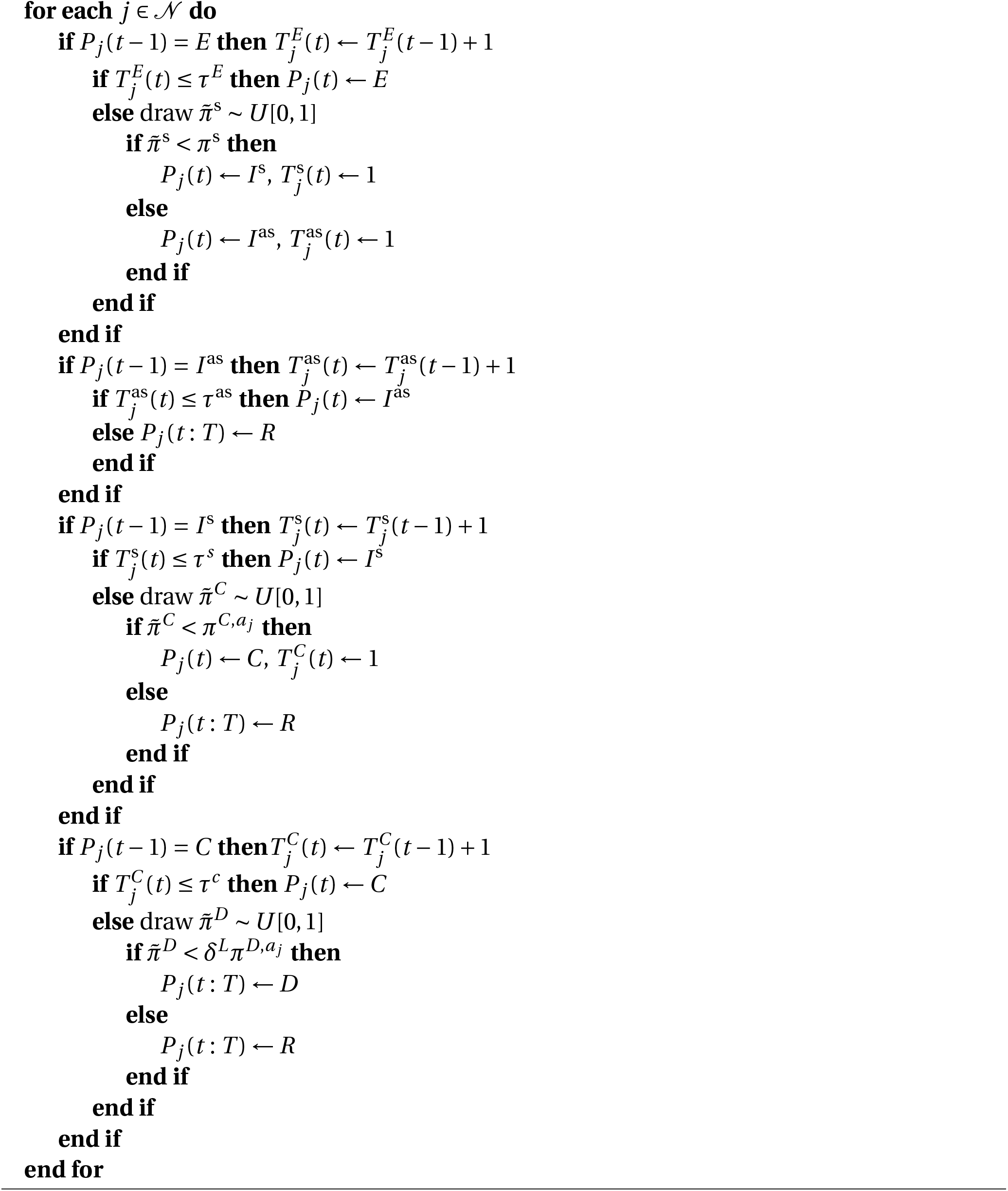

### C.5 Compute New Infections

Next, Algorithm 10 will compute which agents will next become infected by looping over all infected agents and determining how many neighbours they visit and ultimately infect. Infection occurs as a result of two stochastic events in the model: a physical meeting between two agents and the stochastic transmission of the virus conditional on meeting. First, a physical meeting between two agents *i* and *j* occurs with a time-varying, pair-specific probability 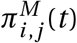 and second, conditional on a physical meeting between an infectious and a susceptible agent, transmission of the disease occurs with probability *π*^*E*^ (the fundamental transmissibility of the virus which we estimated to fit to incidence data). 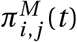 depends on a variety of features within the model. First, agent *j* inevitably has daily physical meetings with all members of their household, i.e. 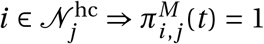. Second, for members in the set of non-household contacts of agent *j*, the probability of a meeting between *j* and 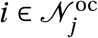 depends on two components: (i) a fundamental parameter that calibrates the likelihood of a meeting between the two agents if they are both fully compliant to lockdown regulations by authorities (*ω* ∈ [0, 1]) and (ii) the degree compliance of *each* agents with lockdown regulations by authorities that aim to prevent transmission. This is modelled as follows: in every period *t*, agent *j* chooses a degree of compliance *φ*_*j*_ (*t*) which is a function of a public signal and observations (via De Groot learning) of the degree of compliance of other agents in the agent’s network as described in Algorithm 8. The probability of a meeting between agent *j* and 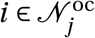 is specified as:

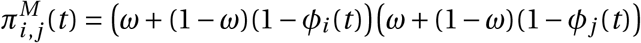

Thus, if both agents are fully compliant (i.e. *φ*_*i*_ (*t*) = *φ*_*j*_ (*t*) = 1), the probability of a physical meeting is *ω*^2^. If both agents are fully non-compliant (i.e. *φ*_*i*_ (*t*) = *φ*_*j*_ (*t*) = 0), the probability of a physical meeting is 1. This encodes two features: (i) even with full compliance to policy, some physical meetings may happen during the course of everyday life, and (ii) individuals can always coordinate to ensure a meeting should they wish to do so strongly enough.

## Appendix D Data Sources

All data used in this paper is publicly available. In this section, we outline the data we used and provide information of how the data can be downloaded.

### Algorithm 10 Compute new infections

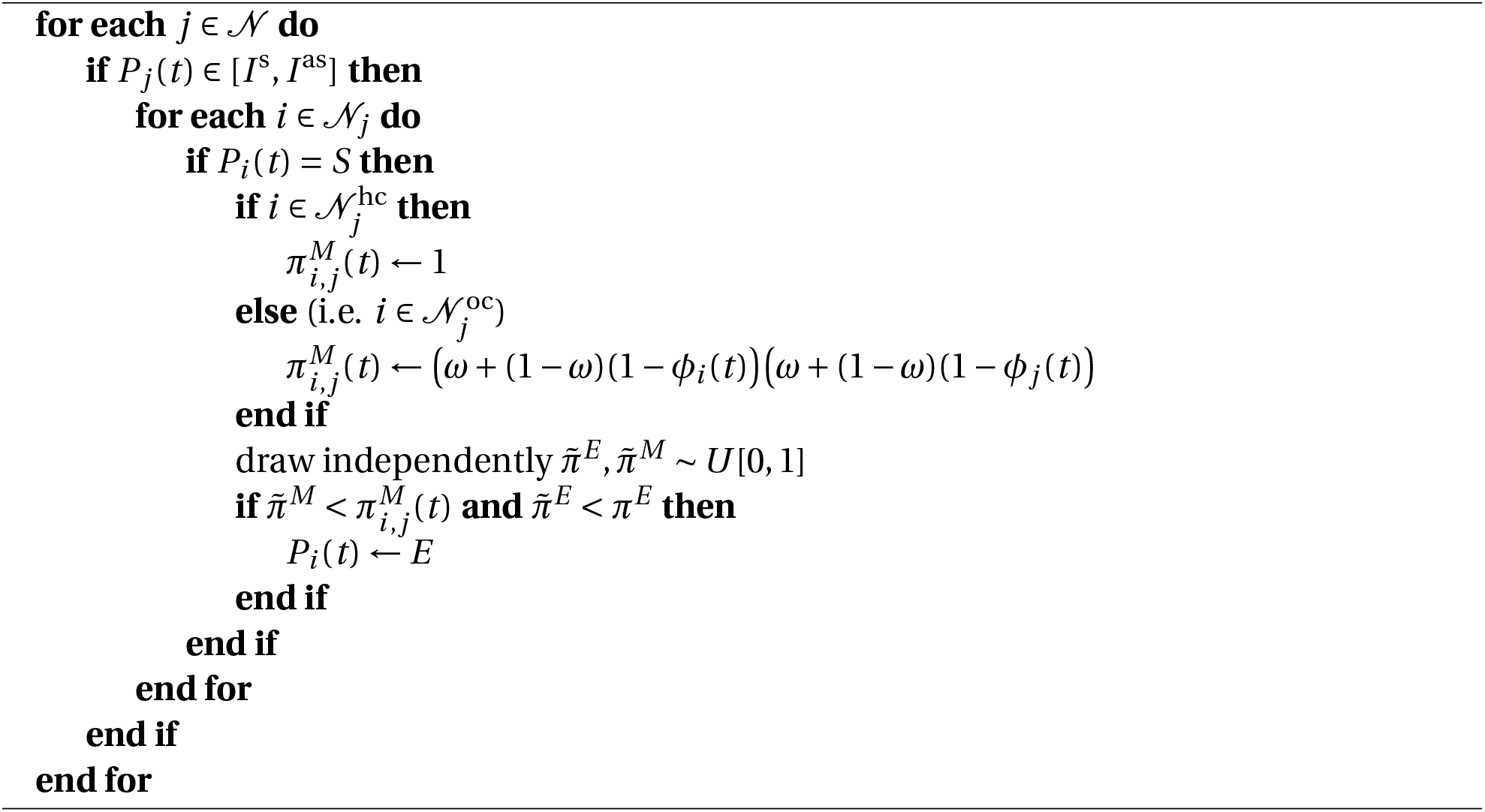

- *F*^*ca*^: Observed cases per district
  Source: Western Cape Government
  Each day, the Western Cape government releases a pdf report documenting the number of cases spatially. These reports can be found here
  We transcribe these cases for each sub-district by hand for each day. A sub-district is a larger spatial definition than the wards we use in this paper.
  We then overlay our wards on the sub-districts and assign wards to sub-districts.
  Finally, we assign sub-district level infection cases to wards based on a probability calculated as the ward level population normalized by the sub-district level population.
- *F*^*hc*^ : Observed age group household contacts
  Source: Prem, Cook and Jit (2017)
  Data can be downloaded from the journal website, here
- *F*^*oc*^ : Observed non-household contacts
  Source: Prem, Cook and Jit (2017)
  Data can be downloaded from the journal website, here
- *F* ^*pop*^ : Observed district population
  Source: 2011 Census from Statistics South Africa
  Data can be downloaded from Statistics South Africa’s website, after the creation of a free profile, here. Once you are logged in, navigate to *Community Profiles* > *Census 2011 (2016 Boundaries)* in the sidebar
- *F* ^*age*^: Observed age distribution per district
  Source: 2011 Census from Statistics South Africa
  Data can be downloaded from Statistics South Africa’s website, after the creation of a free profile, here. Once you are logged in, navigate to *Community Profiles* > *Census 2011 (2016 Boundaries)* in the sidebar
- *F*^*TV*^ : Observed travel matrix
  Source: 2013 National Household Travel Survey from Statistics South Africa
  Data is obtained from DataFirst. Data can be found on the DataFirst data portal, here. You will need to create an account to access the data.
  We discuss the steps taken in mapping this data to our ward spatial structure in Appendix E.
- *F*^*hs*^: Observed district household size distribution
  Source: 2011 Census from Statistics South Africa
  Data can be downloaded from Statistics South Africa’s website, after the creation of a free profile, here. Once you are logged in, navigate to *Community Profiles* > *Census 2011 (2016 Boundaries)* in the sidebar
- *F* ^*in*^: Informality level
  Source: 2011 Census from Statistics South Africa
  Data can be downloaded from Statistics South Africa’s website, after the creation of a free profile, here. Once you are logged in, navigate to *Community Profiles* > *Census 2011 (2016 Boundaries)* in the sidebar
- *F* ^*str in*^: Stringency index
  Source: Oxford Covid-19 Government Response Tracker
  Data can be downloaded from Github, here. Download the OxCGRT_latest.csv file and select country code ZAF and the column StringencyIndex

## Appendix E Travel Calibration

In order to calibrate our model to realistic travel patterns, we use the 2013 National Household Travel Survey, a nationally representative travel survey. Importantly, for our purposes, the survey records where respondents live, where they travel to for work and/or education, the frequency of travel, and the time spent travelling. The travel survey allocates respondents to travel regions of which there are 18 in Cape Town. Our challenge then is to relate the 116 wards we use in this paper, to the 18 travel regions. We illustrate a simplified schematic of how these two structures relate in Figure 9.

**Figure 9:**
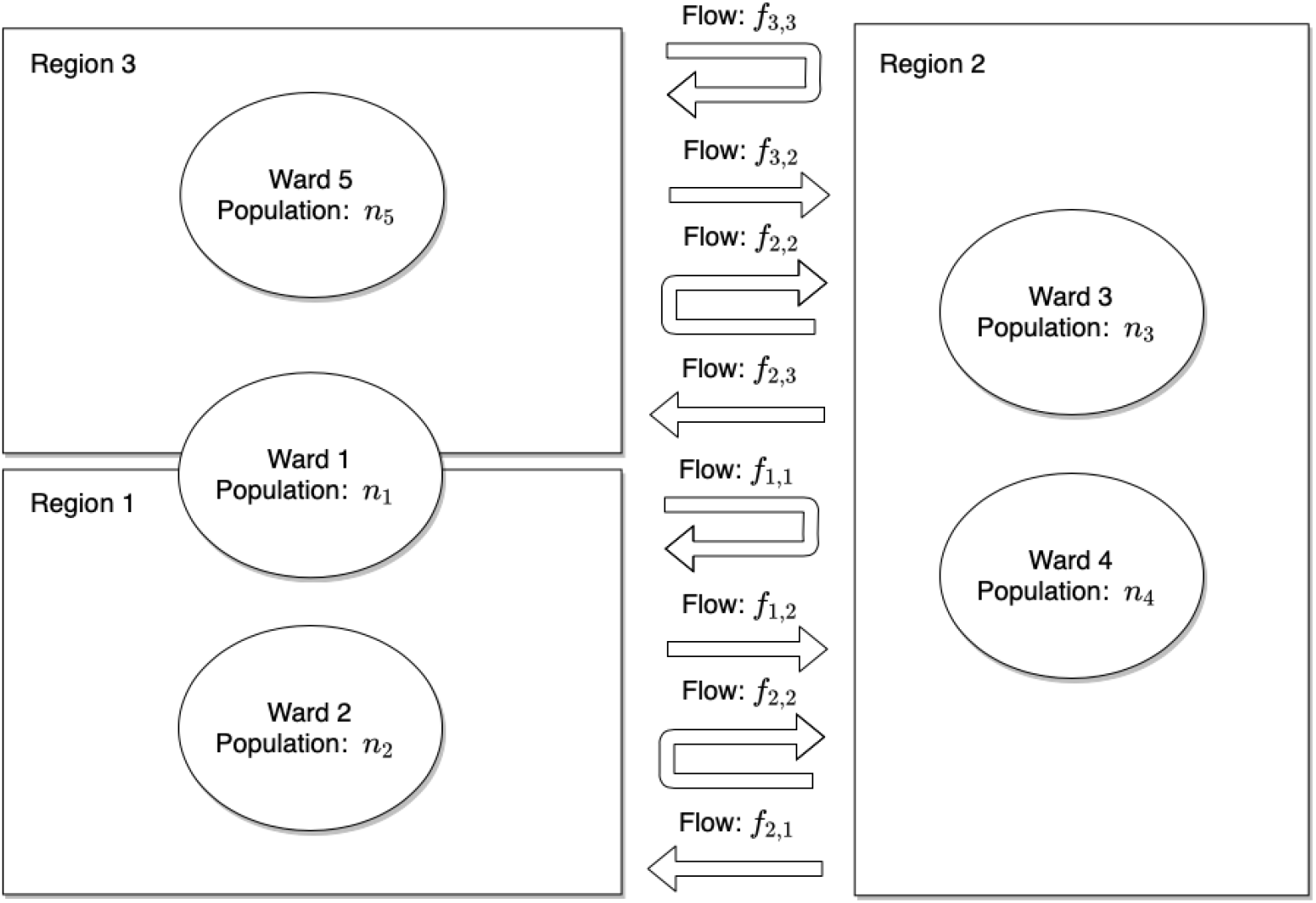
Travel data illustrative schematic. This figure illustrates how the travel survey data we use corresponds to wards, our geo-spatial structure. The travel survey data records flows of people between regions. Respondents are asked in which region they reside and then in which region they commute to for work and/or education reasons. There are 18 regions in Cape Town, while there are 116 wards. In this illustrative schematic, we have 3 regions where travel only occurs between Region 1 and Region 2, and between Region 3 and Region 2. In some cases, wards will fit perfectly into a single region as is the case with Wards 2 through 5. However, there may be a case where a ward overlaps with two regions as is the case with Ward 1 which overlaps Regions 1 and 2.

Our goal is to create a ward-level travel probability matrix using the regional travel data. To implement this, we allocate region level flows to wards in proportion to the size of the both the origin and destination ward population as a share of their respective total regional population. We illustrate this process in Figure 10.

**Figure 10:**
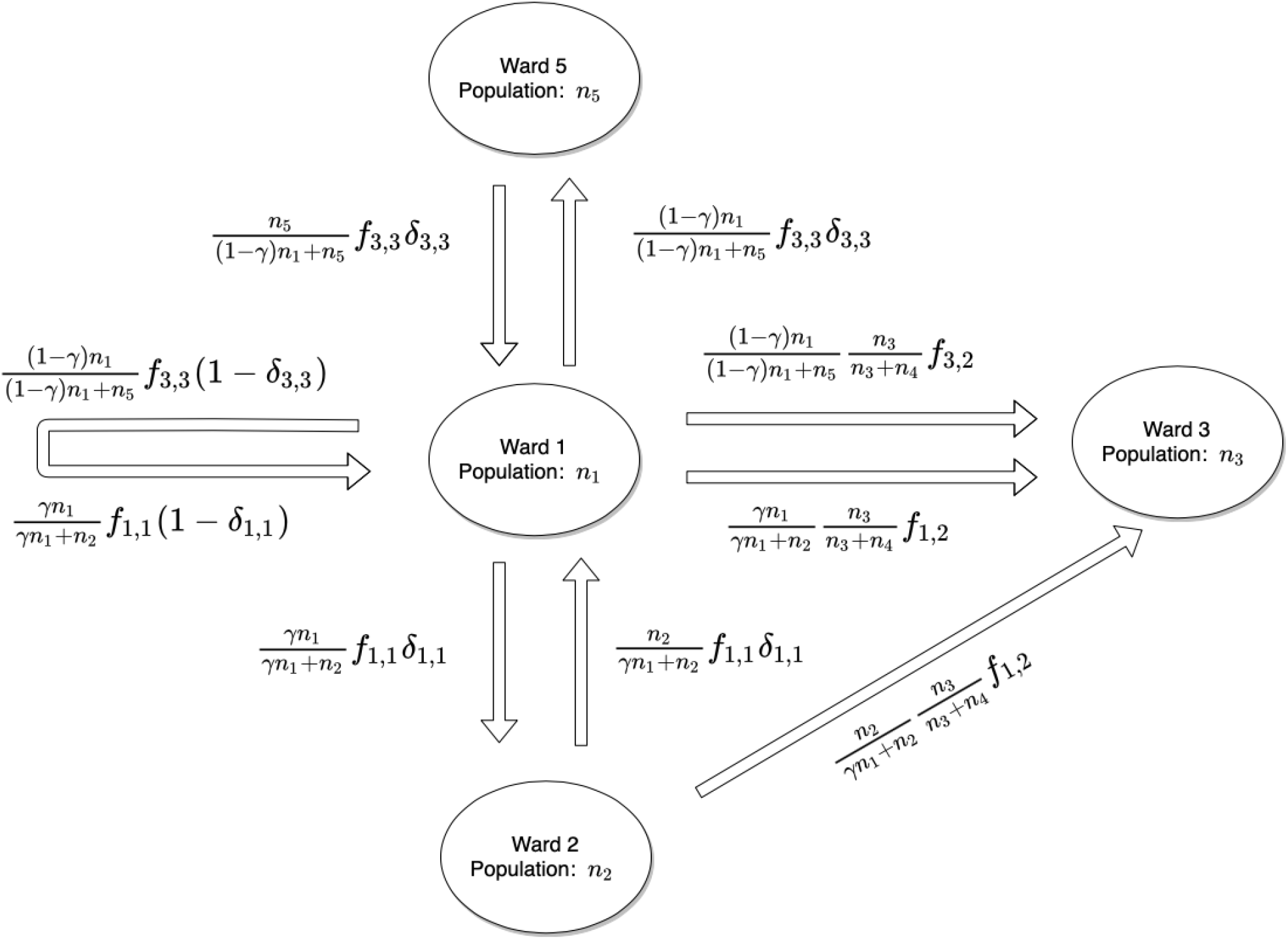
Mapping region-based travel data to ward. This figure illustrates how we map the travel survey data we collect to wards, our suburb structure. We allocate region level flows to wards in proportion to the size of both the origin and destination ward population as a share of their respective total region population. *γ* reflects the percentage overlap between a ward and a region, in the case of an overlap. *f*_*i,j*_ represents flows between regions. *δ*_*i,i*_ represents the share of survey respondents who both live and work / attend school in region *i* and who have an average travel time that is above the 25^th^ percentile of the travel time distribution of all individuals who both live and work / attend school in region *i*.

There are three scenarios which we encounter. The first involves travel between two wards, which are both perfectly located within two different regions, as is the case between Ward 2, located in Region 1, and Ward 3, located in Region 2. We construct the flow of people between these wards as the product of:

- 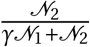: The size of Ward 2’s population relative to the total population of Region 1;
- 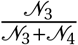: The size of Ward 3’s population relative to the total population of Region 3;
- *f*_1,2_: The flow of people between Region 1 and Region 2;

We introduce a parameter *γ* which scales the population of Ward 1, *𝒩*_1_. This accounts for a scenario where Ward 1 overlaps with two regions, as is illustrated in Figure 9. *γ* then reflects the percentage geographic overlap between Ward 1 and Region 1. We use this geographic overlap to assign the ward population to the respective region.^39^

Our second scenario involves travel between two wards, in which only one ward is perfectly located within a region, as is the case between Ward 1, located in Region 1 and Region 3, and Ward 3, located in Region 2. In this scenario, we map two flows^40^

- 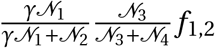: The flow of people from the part of Ward 1 located in Region 1;
- 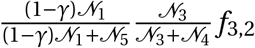: The flow of people from the part of Ward 1 located in Region 3;

Our third scenario represents a case where a respondent reports living and working/attending school in the same region. Within this scenario, we need to decide how to assign respondents who are likely to live and travel within the same ward versus respondents who are likely to live in one ward, but travel to another ward *within the same region*. In our illustrative example, such as case occurs between Ward 1 and Ward 2 and between Ward 1 and Ward 5. In order to allocate these flows, we introduce a new parameter *δ*_*i,i*_ which takes a value between 0 and 1, indicating the likelihood that an individual respondent lives and works / attends schools in different wards within the same region.

We leverage a question in the travel survey, which asks how much time a daily commute takes. Using this question, we take the distribution of travel times for all respondents who live and work/attend school in the same region and assign any individual who has a travel time greater than the 25^th^ percentile of this distribution as a *cross-ward traveller*, assigning the rest of respondents as *within-ward travellers. δ*_*i,i*_ then reflects the share of within region travellers, who are likely to be *cross-ward travellers*. The assumption here being that the likelihood of cross-ward travel increases with travel time. Using this, we now assign 4 travel flows for Ward 1

- 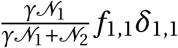: The flow of people from the part of Ward 1 located in Region 1 to Ward 2;
- 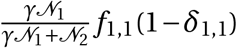: The flow of of people from the part of Ward 1 located in Region 1 who do not travel;
- 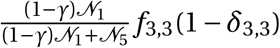: The flow of people from the part of Ward 1 located in Region 3 to Ward 5;
- 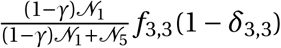:The flow of people from the part of Ward 1 located in Region 3 who do not travel;

Following this approach, we obtain a ward-level travel flow between each ward. To convert this to travel probability, we normalize each outgoing travel flow from a given ward by the total outgoing flows from that same ward. For Ward 1 then, the probability of travelling to Ward 2 can be calculated as follows:

- Flow from Ward 1 to Ward 2: *f*_*w*1,*w*2_;
- Total flows from Ward 1: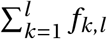;
- Probability of travel between Ward 1 and Ward 2: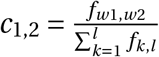;

## Footnotes

1 See, for example, “The simulations driving the world’s response to COVID-19” -Nature, 2 April 2020

2 See Verelst, Willem and Beutels (2016) for a survey of the epidemiological literature that features behavioural change. We discuss the rapidly growing literature of models that feature both disease transmission and some form of optimal agent behaviour in the literature review at the end of this section.

3 See, for example, the National Institute of Communicable Diseases (NICD) in South Africa citing a lack of human behaviour within national epidemiological models as a reason for over-projected deaths: “The known the unknown and the unknowable modelling Covid-19 between scarce data and the need to make decisions”–NICD, 24 July 2020.

4 Based on Hale et al. (2020), who develop a lockdown stringency index which scores lockdowns worldwide on a 0-100 scale where 0 is the least and 100 is the most stringent lockdown. In response to the Covid-19 outbreak, South Africa went into a lockdown with a 87.96 stringency index at the start of our simulation period that was slowly reduced to 80.56 by the end of our period of interest.

5 See, “Model prediction overview from the South African Government”

6 Excess fatalities are generally considered to be more reliable, given widespread concerns about the under-counting of both infections (Adepoju, 2020) and fatalities (Pasquariello and Stranges, 2020; Leon et al., 2020).

7 Testing in South Africa was severely limited at the beginning of the pandemic, hence the number of initial cases cannot be reliably estimated and we treat it as a free parameter.

8 We explicitly focus on the cost of life and do not consider economic costs. For an analysis that incorporates economic cost, see for example Krueger, Uhlig and Xie (2020), Acemoglu et al. (2020) and Eichenbaum, Rebelo and Trabandt (2020).

9 In this context also known as, agent-based, individual-based, or micro-simulation models

10 Our model could be classified as a Susceptible Exposed Infected without symptoms, Infected with symptoms, in need of Critical care, Recovered, Dead model. However like most Covid-19 models—see e.g. Mwalili et al. (2020); Calafiore, Novara and Possieri (2020)—we still classify this as a SIR model because each additional compartment can be seen as a sub-category of these three. We also calibrate our model using contact matrices and detailed population data. We choose the agent-based structure because it naturally incorporates a network structure and using heterogeneous agents means that *local* learning is possible.

11 We sometimes refer to physical interactions as social connections.

12 In Appendix B, we provide a detailed pseudo-code description of the model.

13 We use the convention that sets are denoted by formal script, e.g. *𝒩* ; the value of a variable (such as the cardinality of a set) are denoted by upper case letters (e.g. *N* ≡ |*𝒩* |), and generic elements are denoted by lower case letters (e.g. an agent is denoted as *j* ∈ *𝒩*). Indices denoting generic elements of a set are subscripts, while additional identifiers are superscripts.

14 The epidemiological literature denotes these values as *compartments* and we follow this nomenclature on occasion.

15 We use this simplifying assumption to model the situation where patients who are isolated in hospital cannot spread the disease in their regular social networks.

16 *F* ^hc^is a matrix that records the best estimates of data on daily number of contacts between individuals in different age categories within the same household. It has the following structure: row *a* of the square matrix *F* ^hc^contains the list of average daily number of contacts that an individual in age category *a* is expected to have with an individual in each of the age categories represented by the columns of *F* ^hc^. A probability distribution of likely contact of someone of age category *a* with someone of age category *b* is constructed by normalizing the entries of row *a* of *F* ^hc^so that they add to one.

17 We observe the empirical distribution of households sizes for each district using national census data. Since the algorithm is stochastic and we use representative agent populations smaller than the actual populations in the city modelled, there are some additional algorithmic features that ensure that (i) the maximum household size randomly drawn remains smaller than remaining number of agents to be assigned to a household at every point in the algorithm, and (ii) that the final number of agents in a district correspond to the proportional size of that district given the ratio of total modeled agents to actual population of the city.

18 Its structure is identical to that of *F* ^hc^described in footnote 16.

19 See Appendix E for the details on the construction of *F* ^tv^.

20 The algorithm is robust to implementations with smaller populations where it is possible that 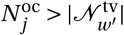. See Algorithm 4.

21 ^21^The basic infectiousness of the disease *π*^*E*^ is an uncertain parameter that we treat as a constant over time and across individuals in this model as a simplification (see Section 3.2 for our calibration.). As more certain results become available in the medical literature, simple extensions of the model can consider situations where e.g. children are less likely to become infected and/or transmit the disease than older individuals.

22 See footnote 21 for a discussion on the basic infectiousness of the disease *π*^*E*^, which is an uncertain parameter.

23 ^23^Put differently, *ω*^2^ represents a floor on the level of mobility.

24 Economic epidemiological models encode a similar logic whereby agents trade off wanting to leave home to earn an income to satisfy their economic needs versus wanting to stay at home to minimize the risk of infection. In these models, no matter how high the infection cost is, there will always be agents whereby the economic benefit from making an contact with others exceed the cost of infection, given a distribution of economic needs. The forces that drive these kinds of agents to always make trips, irrespective of infection risk, are captured in *ω*.

25 Find the code and replication files on our GitHub repository, here.

26 *T =* 1, 171 is the length of our empirical reference data.

27 There likely is no general transmission probability when two people meet since local factors such as climate, ventilation, and cultural proximity affect this parameter.

28 The latest date for which we have survey data is 2011.

29 Hence, we will refer to them as wards from here on out.

30 The travel survey only asks respondents where they live, work and where they attend school. As a result, travel patterns reflect patterns related specifically to work and school and not for other reasons such as leisure or shopping, for example.

31 See for example: “Winde confirms pressure on hospital system increased, despite not yet hitting peak capacity” -News24, 22 May 2020

32 Available to download, here. Accessed on 04/17/2021.

33 Hale et al. (2020) develop a lockdown stringency index which scores lockdowns worldwide on a 0-100 scale where 0 is the least and 100 is the most stringent lockdown. In response to the Covid-19 outbreak, South Africa went into a 87.96 stringency index lockdown, one of the strictest lockdowns in the world, enforced by a large police and army presence in the streets.

34 The seminal papers developing SMM are McFadden (1989); Duffie and Singleton (1990); Lee and Ingram (1991).

35 The constraint restricts input parameters to positive values only and was developed by Alex Blaessle, source code is available here.

36 See the South African Medical Research Council’s website for more information regarding data cleaning and statistical methods used.

37 Computations were performed using facilities provided by the University of Cape Town’s ICTS High Performance Computing team (hpc.uct.ac.za) and the University of Stellenbosch’s HPC1, Rhasatsha (www.sun.ac.za/hpc).

38 Thompson (2021); Voysey et al. (2021) present evidence that the vast majority of vaccinated individuals will both become immune to the effects of the virus and will no longer transmit it.

39 The inherent assumption here being that the ward level population is distributed evenly across the ward.

40 In the event that the destination ward also overlaps multiple regions, we can modify these flow equations with an additional overlap parameter which scales the destination wards population

